# Latent class analysis of symptoms across schizophrenia, schizoaffective disorder, and bipolar I disorder

**DOI:** 10.1101/2025.10.31.25339091

**Authors:** Dana M. Lapato, Roseann E. Peterson, Eric D. Achtyes, Peter F. Buckley, Ayman H. Fanous, Douglas S. Lehrer, Humberto Nicolini, Dolores Malaspina, Mark H. Rapaport, GPC Investigators, Kenneth S. Kendler, Michele Pato, Carlos Pato, Tim B. Bigdeli

## Abstract

Data-driven phenotypes have the potential to accelerate biomedical research but must be vetted thoroughly for robustness, interpretability, and generalizability. This study sought to create and evaluate the predictive validity of empirical phenotypes derived from assessments of signs and symptoms of psychopathology collected in a large cohort of patients using a validated semi-structured clinical interview based on the OPCRIT system. Data was available for N = 17,719 individuals diagnosed with schizophrenia (n = 10,429; 30% female), bipolar I disorder (n = 4,520; 53% female), or schizoaffective disorder (n = 2,770; 44% female) using DSM-IV-TR criteria. The best-fitting latent class analysis (LCA) model was a 6-class solution. Three of the six class profiles replicated results from previous studies. Stratifying by sex did not alter class profiles. Latent classes were significantly associated with established DSM diagnoses for primary and sex-stratified results but not for the LCA solution based primarily on symptoms and lacking indicators related to illness course and relative prevalence of psychotic versus affective symptoms in the overall clinical presentation. All LCAs (i.e., primary, sex-stratified, and symptom-only analyses) produced latent classes that showed statistically significant associations with demographic, etiological, and clinical correlates, but the effect sizes were modest for most associations and comparable in magnitude to those observed for established DSM diagnosis. These results collectively suggest that symptom-based empirical phenotypes derived from LCA may not outperform DSM diagnoses and reaffirm the long-observed importance of illness course for differentiating schizophrenia and bipolar I disorder.

## Introduction

Despite a substantial amount of effort to characterize the nosology of psychotic disorders^1–4^, uncertainty remains regarding the optimal classification structure. Much of the modern conceptualization of psychotic disorder nosology hails from Kraepelin’s descriptions of dementia praecox (later termed “schizophrenia” by Bleuler) and manic-depressive insanity^1^. Over the last thirty years, both data-driven and dimensional approaches have been used to interrogate the boundaries of nonaffective and affective psychopathology and identify more homogeneous subpopulations within and across the diagnoses of schizophrenia (SCZ), schizoaffective disorder (SAD), and bipolar I disorder (BD1)^2–7^. However, extending these phenotyping results to inform and improve gene discovery in genome-wide association studies of schizophrenia and other disorders has been challenging, especially regarding the empirical phenotypes derived from latent class analysis (LCA).

LCA is a person-centered statistical approach that assigns participants to mutually exclusive groups based on the similarity of responses to a collection of indicators. The method is intuitively appealing for psychiatric genetics phenotyping because its clustering process is analogous to clinical nosology approaches and results in discrete groups of patients and class profiles that can be more easily interpretable and readily comparable to DSM-based diagnoses compared to dimensional phenotypes (i.e., factor scores). Thus far, there have been four LCAs of psychotic disorders that included at least 300 participants, collected symptom-level measures, and enrolled a multi-diagnosis cohort that included at least schizophrenia and bipolar I disorder ^5,8–10^.

Across these four studies, several commonalities in the latent class symptom profiles have emerged despite marked differences in participant demographics, clinical characteristics, psychometric instruments, and specific indicators included in the models. More recent work by the Bipolar-Schizophrenia Network on Intermediate Phenotyping (B-SNIP) has extended the use of LCA to cluster patients by neurobiological measures and identify potential novel psychosis biotypes^11–14^.

While these results are exciting, the extent that existing LCA results have been applied to large-scale genetic studies is limited. In the case of the psychosis biotypes, replication is challenging because almost no study has collected the diverse array of measures needed to assign participants to one of the three psychosis biotypes. For the symptom-based LCA studies, reusing the derived classes is risky because the generalizability and replicability of the proposed classes have not been evaluated. This limitation is substantial because LCA results can be strongly influenced by dataset-specific characteristics (e.g., a SCZ cohort comprised entirely of adolescent patients who have received in-patient care may yield different latent class profiles than a SCZ cohort of older adults who have never been hospitalized). Thus, there remains a need to conduct a large-scale empirical phenotyping study of the signs and symptoms of psychosis in a cohort diverse enough to enable the identification of potentially rare subgroups and large enough to evaluate the robustness and generalizability of the results by sex. To that end, we conducted LCA in the largest transdiagnostic case-only cohort of psychotic illness to date and compared the predictive validity of the data-driven phenotypes to DSM-based diagnoses. We also conducted stratified analyses to assess the impact of sex on both the symptom profiles and association strength of class assignment to demographic, etiological, and clinical measures. Finally, we reran the full cohort LCA using primarily symptom-level indicators (i.e., no indicators that represent illness course or the relative prevalence of psychotic versus affective symptoms in the clinical picture) and assessed the impact on latent class number, composition, and correlates. The purpose of this sensitivity analysis was to recapitulate some of the symptom-focused phenotyping approaches that have been employed in GWAS and to evaluate the utility of LCA for deriving empirical clusters in situations where information about illness course is absent.

## Methods

### Sample

The cohort analyzed in this study came from Genomic Psychiatry Cohort^15^ participants diagnosed with schizophrenia, schizoaffective disorder, or bipolar I disorder. The GPC recruited participants at more than a dozen clinical sites and across a variety of settings, including acute and chronic care inpatient facilities, outpatient facilities, and community settings. Institutional review board approval was received for all sites, and informed consent was obtained from all participants.

### Diagnostic assessment

All participants enrolled as probable cases were interviewed by trained mental health professionals and research staff using the Diagnostic Interview for Psychosis and Affective Disorders (DI-PAD). Clinicians confirmed participants’ established diagnosis for schizophrenia, schizoaffective disorder, or bipolar disorder based on criteria from the fourth edition of the Diagnostic and Statistical Manual of Mental Disorders (DSM-IV-TR^16^). Where possible, information about disorder subtype was noted from medical records or participant recall. The DI-PAD is a semi-structured clinical interview based on the Diagnostic Interview for Genetic Studies (DIGS^17^) and is designed to capture phenotypic heterogeneity in affective and nonaffective psychotic illnesses. The DI-PAD includes a mixture of free response, Likert-type, and binary items from the Operational Criteria Checklist (OPCRIT^18^) that collectively assesses a broad array of signs and symptoms of psychopathology, clinical characteristics, risk factors, current and premorbid functioning, and illness course.

No formal studies of the DI-PAD inter-rater reliability across GPC sites have been published, but the GPC staff conduct inter-rater and inter-site reliability tests annually during site visits. These assessments include study visit observations and side-by-side ratings between site interviewers and visiting trainers. Separate studies evaluating the reliability of OPCRIT-based phenotypes have reported that OPCRIT diagnoses generally have good to excellent agreement with the best-estimate lifetime consensus procedure and ICD-10 codes^19,20^ and substantial inter-rater reliability for both individual and diagnostic-level items^21^.

### Exclusion criteria

Any individual who self-reported or had medical records indicating the presence of a coarse brain disorder or other physical illness (e.g., overt brain lesions or marked metabolic disturbances known to cause psychotic symptoms) prior to psychiatric symptom onset that could explain most or all the current psychiatric symptoms was excluded from this analysis (OPCRIT 15). Individuals diagnosed with bipolar I disorder were not required to endorse psychotic symptoms; however, bipolar I disorder cases with psychosis were intentionally oversampled compared to nonpsychotic bipolar I disorder. Individuals diagnosed with bipolar II disorder were excluded from this study.

### Data analysis

The poLCA R package was used to conduct the latent class analysis^22,23^ (see supplemental file for additional details about the coding environment). The primary analysis included the entire cohort and modeled 19 indicators based on 28 symptom-level OPCRIT items that had been recoded using a similar algorithm as prior studies (see Table S1)^5,10^. The second analysis used the same 19 indicators applied to male and female cohorts separately to evaluate the generalizability of the LCA across sexes. The third analysis was identical to the primary analysis except that it included primarily symptom indicators (i.e., did not include the two indicators that captured illness course [OPCRIT 90] and the relative prevalence of psychotic versus affective symptoms [OPCRIT 52]).

The same procedure for latent class extraction was followed for each analysis. First, starting with a two-class solution, additional latent classes were extracted until at least one class was smaller than 6% of the sample. Each model was refitted 30 times with random starting values and a maximum of 5000 iterations per run to address potential issues with local maxima and nonconvergence. Fit statistics and posterior probabilities quantifying the likelihood that a participant belonged in each class were recorded for the best-fitting solution for each class solution (i.e., 2-class solution, 3-class solution). Next, class solutions were examined for interpretability and overall model fit. Similar to other LCA studies of psychotic illness^9,10^, model selection was guided by the examination of the log likelihood (−2LL), Akaike Information Criterion (AIC^24^), Bayesian Information Criterion (BIC^25^), class size, and the interpretability of the class structures. Such a holistic evaluation is common for LCAs because purely empirical approaches rarely produce optimal results in terms of interpretability and external validation and because no individual metric or method for determining the best number of classes to extract has universal endorsement^26^. For each analysis, the external validity of the latent class assignments from the overall best fitting model was evaluated using demographic, etiological, and clinical characteristic measures. Effect sizes are provided in addition to p-values to facilitate interpretability given that the large cohort size made achieving statistical significance using traditional thresholds trivial. Effect sizes were calculated using either eta squared (η^2^) or Cohen’s omega (ω) for ANOVA and chi square, respectively^27^. Thresholds defining small (η^2^ < 0.06; ω < 0.30), medium (0.06 < η^2^ < 0.14; 0.30 < ω < 0.50), and large (η^2^ >= 0.14; ω >= 0.50) effects were established in Cohen (1988)^28^.

## Results

Data was available for N = 17,719 individuals with a diagnosis of SCZ (n = 10,429), BD1 (n = 4,520), or SAD (n = 2,770). The sex-specific LCA models were applied to 10,780 male (SCZ = 7,122; BD1 = 2,111; SAD = 1,547) and 6,628 female (SCZ = 3,027; BD1 = 2,409; SAD = 1,192) participants. Cohort demographics are presented in Table 1. The majority of participants in this study were male (61%), interviewed between 30-50 years old, and reported an age of onset in their early twenties. We observed statistically significant differences across diagnostic groups with respect to various demographic, etiologic, and clinical characteristics; most effect sizes were small (η^2^ = 0.0022 – 0.03).

**Table 1.** Cohort Demographics.

| Characteristic | Overall N =<br>17,719<sup>1</sup> | Schizophrenia, N =<br>10429 (59%)<sup>1</sup> | Bipolar I, N =<br>4520 (26%)<sup>1</sup> | Schizoaffective, N =<br>2770 (16%)<sup>1</sup> | p-value<sup>2</sup> |
| --- | --- | --- | --- | --- | --- |
| **Age of Onset** | 20.5 (8.7) | 21.4 (8.2) | 19.6 (9.2) | 18.7 (8.7) | **<0.0005** |
| **Sex** |  |  |  |  | **<0.0005** |
| Female | 6,628 /<br>17408<br>(38%) | 3,027 / 10149 (30%) | 2,409 / 4520<br>(53%) | 1,192 / 2739 (44%) |  |
| Male | 10,780 /<br>17408<br>(62%) | 7,122 / 10149 (70%) | 2,111 / 4520<br>(47%) | 1,547 / 2739 (56%) |  |
| **Family History (SCZ)** |  |  |  |  | **<0.0005** |
| Family history of schizophrenia | 4,798 /<br>15994<br>(30%) | 2,553 / 8843 (29%) | 1,192 / 4455<br>(27%) | 1,053 / 2696 (39%) |  |
| No family history of schizophrenia | 11,196 /<br>15994<br>(70%) | 6,290 / 8843 (71%) | 3,263 / 4455<br>(73%) | 1,643 / 2696 (61%) |  |
| **Family History (other)** |  |  |  |  | **<0.0005** |
| Family history of psychiatric disorder other than schizophrenia | 6,962 /<br>15981<br>(44%) | 2,992 / 8832 (34%) | 2,578 / 4460<br>(58%) | 1,392 / 2689 (52%) |  |
| No family history | 9,019 /<br>15981<br>(56%) | 5,840 / 8832 (66%) | 1,882 / 4460<br>(42%) | 1,297 / 2689 (48%) |  |
| **Stressors at Onset** |  |  |  |  | **<0.0005** |
| None present | 9,987 /<br>16010<br>(62%) | 6,326 / 8849 (71%) | 2,298 / 4461<br>(52%) | 1,363 / 2700 (50%) |  |
| Present | 6,023 /<br>16010<br>(38%) | 2,523 / 8849 (29%) | 2,163 / 4461<br>(48%) | 1,337 / 2700 (50%) |  |
| **Course of Disorder** |  |  |  |  | **<0.0005** |
| 1. Single episode with good recovery | 52 / 15988<br>(0.3%) | 20 / 8846 (0.2%) | 31 / 4454<br>(0.7%) | 1 / 2688 (<0.1%) |  |
| 2. Multiple episodes with good recovery between | 532 / 15988<br>(3.3%) | 13 / 8846 (0.1%) | 509 / 4454<br>(11%) | 10 / 2688 (0.4%) |  |
| 3. Multiple episodes with partial recovery between | 3,117 /<br>15988<br>(19%) | 36 / 8846 (0.4%) | 2,850 / 4454<br>(64%) | 231 / 2688 (8.6%) |  |
| 4. Gradual onset over period up to six months | 7,222 /<br>15988<br>(45%) | 4,823 / 8846 (55%) | 821 / 4454<br>(18%) | 1,578 / 2688 (59%) |  |
| 5. Insidious onset over period greater than six months | 5,065 /<br>15988<br>(32%) | 3,954 / 8846 (45%) | 243 / 4454<br>(5.5%) | 868 / 2688 (32%) |  |
| **Self-identified Race** |  |  |  |  | **<0.0005** |
| African American Or Black | 6,859 / 17717 (39%) | 4,049 / 10428 (39%) | 1,579 / 4520 (35%) | 1,231 / 2769 (44%) |  |
| American Indian/Alaskan Native | 249 / 17717 (1.4%) | 141 / 10428 (1.4%) | 50 / 4520 (1.1%) | 58 / 2769 (2.1%) |  |
| Asian | 301 / 17717 (1.7%) | 200 / 10428 (1.9%) | 58 / 4520 (1.3%) | 43 / 2769 (1.6%) |  |
| More Than One Race | 1,441 / 17717 (8.1%) | 710 / 10428 (6.8%) | 455 / 4520 (10%) | 276 / 2769 (10.0%) |  |
| Native Hawaiian/Pacific Islander | 59 / 17717 (0.3%) | 36 / 10428 (0.3%) | 13 / 4520 (0.3%) | 10 / 2769 (0.4%) |  |
| Other/Unknown | 1,399 / 17717 (7.9%) | 818 / 10428 (7.8%) | 376 / 4520 (8.3%) | 205 / 2769 (7.4%) |  |
| White | 7,409 / 17717 (42%) | 4,474 / 10428 (43%) | 1,989 / 4520 (44%) | 946 / 2769 (34%) |  |

<sup>1</sup>Mean (SD); n / N (%)
<sup>2</sup>Kruskal-Wallis rank sum test; Pearson's Chi-squared test. For Pearson's Chi-squared tests, p-values were calculated via Monte Carlo simulation (n=2000 replicates).

There were two exceptions to this pattern. First, individuals who endorsed having a first or second degree relative diagnosed with a psychiatric disorder were more likely to have a diagnosis of SAD or BD1 compared to SCZ (η^2^ = 0.22, p < 0.0005). Second, a significantly larger proportion of individuals reporting at least one significant stressor present at disorder onset were diagnosed with BD1(48%) or SAD (50%) compared to SCZ (29%; η^2^ = 0.21, p < 0.0005).

### Latent Class Differences in Item Prevalence

For the primary analysis, models with two through six latent classes were tested. Model fit improved as class number increased with the 6-class solution having the best overall model fit indices (Table S2) and interpretability. LCA solution stability was assessed by increasing the number of iterations from 30 to 2000, which produced nearly identical results. Endorsement patterns for most of 19 indicators showed marked differences in endorsement frequencies by class assignment for the 6-class LCA solution (Table S3). Two of the most strongly differentiating indicators across latent classes were *Course of Illness* (OPCRIT 90) and *Psychotic Versus Affective Symptoms* (OPCRIT 52). These two indicators showed a strong relationship not only with latent class membership but also with established diagnosis (Table S4, Table S5). The two weakest indicators for differentiating latent class membership were *Positive Thought Disorder* (endorsement range: 5.5% to 49.3%) and *Deterioration from Premorbid Condition* (endorsement range: 88.9% to 100%).

Overall, the best-fitting LCA solutions for the male-only and female-only sensitivity analyses strongly resembled the primary analysis in terms of item endorsement patterns across classes (fit indices Table S6, Table S7; item endorsement Table S8, Table S9). The main difference between the symptom profiles derived from the full cohort and sex-specific analyses was that the female-only analysis included a symptom profile for schizophrenia with mood swings but not a classic schizophrenia symptom profile lacking affective symptoms (e.g., depressed mood).

The sensitivity analysis without illness course indicators produced latent classes with noticeably weaker inter-class separation by item endorsement pattern not only in the best fitting model (Table S10, Table S11), but also in the other solutions. Neither reducing the number of extracted classes to four classes nor extending it to eight noticeably improved the symptom profile distinctiveness, suggesting that the poor class separation was not an artifact of number of classes.

### Latent Class Descriptions for the Best Fitting LCA for the Primary Analysis

Class 6 was the largest class (22.0% of the sample). This class was marked by prominent delusions, hallucinations, and negative symptoms with little to no affective disturbances and a chronic course of illness. Symptoms related to mania, depression, and psychomotor changes were largely absent. Overall, this class contained the highest level of impaired insight (37%) and the greatest proportion of male participants (74%). This symptom profile resembles classical schizophrenia (“SCZ”).

Class 1 was the next largest class, comprising 20.2% of the sample. This class was marked by high endorsement rates of auditory hallucinations, persecutory delusions, low-to-moderate levels of symptoms associated with mood, appetite/weight, and psychomotor changes, and a chronic course of illness. Affective symptoms played a relatively minor role in the clinical presentation of this class. A clinical picture consisting of prominent psychotic symptoms with intermittent affective disturbances is consistent with schizophrenia with moderate mood swings (“SCZ-Mood”).

Class 5 was the third largest class at 18.4% of the sample. Its symptom profile broadly resembled Class 6 in the relative rates of psychotic symptoms but with much higher rates of depressive symptoms. In both classes, psychotic symptoms, chronic illness, and negative symptoms dominate the clinical picture. The male-to-female ratio for this class resembles the overall cohort demographics (i.e., slightly male-enriched). We will refer to this class as schizoaffective disorder depressed subtype (“SAD-Dep”).

Class 2 (17.3% of sample) was notable for having high levels of auditory hallucinations and manic symptoms as part of an overall clinical presentation predominated by affective symptoms. Positive thought disorder, negative symptoms, and delusions were present but not highly prevalent. While deterioration from premorbid condition was common, most of the individuals in this class experienced episodic illness with good-to-partial recovery in between episodes. This class was one of only two with an excess of female participants (52% versus 39% in the full cohort). This profile is consistent with bipolar I disorder with psychosis (“Psychotic Bipolar”).

Individuals in Class 3 (16.2%) endorsed the broadest array of symptoms, including high endorsement rates of psychotic, manic, and depressive symptoms. Participants in this class had the greatest proportion of individuals classified as presenting psychotic and affective symptoms equally. Affective symptoms predominated in a small minority of participants (∼7%), and psychotic symptoms predominated in ∼21% of class members. Overall, the levels of negative and positive symptoms were moderate and the proportion of female-to-male participants was roughly similar (43% versus 57%, respectively). No single term concisely but comprehensively captures this class. We will refer to it as schizoaffective disorder bipolar type (“SAD-Bip”).

Class 4 was the smallest by far, comprising only 5.8% of the sample. This class had both the highest proportion of female participants (57% versus 39% in the overall cohort) and the lowest prevalence of psychotic symptoms. None of the individuals assigned to Class 4 endorsed Schneiderian delusions or hallucinations, and only three percent of individuals had any psychotic symptoms present. Both manic and depressive symptoms were prevalent and noted as appearing absent psychotic symptoms. The predominant course of illness was episodic with good to partial recovery in between episodes. Overall, this clinical presentation resembles a milder BD1 presentation compared to Class 2, and we will refer to it as “Nonpsychotic Bipolar”.

The clinical presentation descriptions from the primary analysis generally apply to sex-specific analyses, although the relative size of the different classes varied compared to the full cohort results. This observation was unsurprising given that the item endorsement patterns for the analyses were relatively similar. In contrast, the latent classes from the LCA solution lacking course of illness indicators proved more challenging to interpret. The class profiles produced from the symptom-focused LCA were reminiscent of the primary results but less distinctive. As a result, the class names used in the tables are similar but not identical to the labels used in the primary and sex-stratified analyses.

### Comparison of Established Diagnoses with Predicted Class Membership

Predicted latent class membership was strongly associated with DSM-IV-TR diagnoses for the primary analysis (Table 2) but was not associated with schizophrenia subtype (i.e., disorganized, paranoid, undifferentiated). Classes 2, 4, and 6 were composed almost exclusively of one diagnosis group. Classes 1, 3, and 5 showed more heterogeneity; each had one diagnosis associated with the majority of class members and had between 13-32% of remaining members split between the other two diagnoses. This pattern largely held for the sex-specific sensitivity analyses (Table S12, Table S13) but was severely attenuated in the second sensitivity analysis which lacked the two course of illness indicators (OPCRIT 90, OPCRIT 52; Table 3). Indeed, the LCA without illness course indicators was the only solution where substantial proportions of both schizophrenia and bipolar I disorder participants were assigned to the same classes and where a majority of the classes (four out of six) showed significant diagnostic heterogeneity.

**Table 2.** Relationship Between Established Diagnosis and Latent Class Assignment Using All Indicators.

| Characteristic | Overall<br>N =<br>17,719<sup>1</sup> | Class 1:<br>SCZ-Mood<br>N =<br>3584<br>(20%)<sup>1</sup> | Class 2:<br>Psychotic<br>Bipolar<br>N = 3073<br>(17%)<sup>1</sup> | Class 3:<br>SAD-Bip<br>N =<br>2863<br>(16%)<sup>1</sup> | Class 4:<br>Nonpsychotic<br>Bipolar<br>N = 1036<br>(5.8%)<sup>1</sup> | Class 5:<br>SAD-Dep<br>N = 3262<br>(18%)<sup>1</sup> | Class 6:<br>SCZ<br>N = 3901<br>(22%)<sup>1</sup> | p-value* | Cohen's<br>w |
| --- | --- | --- | --- | --- | --- | --- | --- | --- | --- |
| **Dx** |  |  |  |  |  |  |  | **<0.0005** | **1.13** |
| Schizophrenia | 10,429 /<br>17719<br>(59%) | **3,096 /**<br>**3584**<br>**(86%)** | 10 / 3073<br>(0.3%) | **605 /**<br>**2863**<br>**(21%)** | 3 / 1036 (0.3%) | 2,827 /<br>3262<br>(87%) | 3,888 /<br>3901<br>(100%) |  |  |
| Bipolar I | 4,520 /<br>17719<br>(26%) | **118 /**<br>**3584**<br>**(3.3%)** | 3,042 /<br>3073 (99%) | **326 /**<br>**2863**<br>**(11%)** | 1,032 / 1036<br>(100%) | 1 / 3262<br>(<0.1%) | 1 / 3901<br>(<0.1%) |  |  |
| Schizoaffective | 2,770 /<br>17719<br>(16%) | **370 /**<br>**3584**<br>**(10%)** | 21 / 3073<br>(0.7%) | **1,932 /**<br>**2863**<br>**(67%)** | 1 / 1036<br>(<0.1%) | 434 /<br>3262<br>(13%) | 12 /<br>3901<br>(0.3%) |  |  |

<sup>1</sup> n / N (%)
\* Pearson's Chi-squared test; p-values were calculated via Monte Carlo simulation (n=2000 replicates).
Bolded columns indicate classes with high diagnostic heterogeneity.
SCZ-Mood = schizophrenia with moderate mood swings; SAD-Bip = schizoaffective bipolar subtype; SAD-Dep = schizoaffective depressed subtype; SCZ = schizophrenia

**Table 3.** Relationship Between Established Diagnosis and Latent Class Assignment Without Illness Course Indicators.

| Characteristic | Overall N =<br>17,719<sup>1</sup> | Class 1:<br>Bipolar<br>N = 4505<br>(25%)<sup>1</sup> | Class 2:<br>SCZ<br>N = 3820<br>(22%)<sup>1</sup> | Class 3:<br>SCZ-Mood<br>N = 2785<br>(16%)<sup>1</sup> | Class 4:<br>SAD-Dep<br>N = 1935<br>(11%)<sup>1</sup> | Class 5:<br>Bipolar-Severe<br>Mania<br>N = 1558<br>(8.8%)<sup>1</sup> | Class 6:<br>SAD-Bip<br>N = 3116<br>(18%)<sup>1</sup> | p-value* | Cohen's<br>w |
| --- | --- | --- | --- | --- | --- | --- | --- | --- | --- |
| **Dx** |  |  |  |  |  |  |  | **<0.0005** | **0.76** |
| Schizophrenia | 10,429 /<br>17719<br>(59%) | **1,603 /**<br>**4505**<br>**(36%)** | 3,803 /<br>3820<br>(100%) | 2,489 /<br>2785<br>(89%) | **1,120 /**<br>**1935**<br>**(58%)** | **1,046 / 1558**<br>**(67%)** | **368 /**<br>**3116**<br>**(12%)** |  |  |
| Bipolar I | 4,520 /<br>17719<br>(26%) | **2,645 /**<br>**4505**<br>**(59%)** | 1 / 3820<br>(<0.1%) | 16 / 2785<br>(0.6%) | **362 /**<br>**1935**<br>**(19%)** | **305 / 1558**<br>**(20%)** | **1,191 /**<br>**3116**<br>**(38%)** |  |  |
| Schizoaffective | 2,770 /<br>17719<br>(16%) | **257 / 4505**<br>**(5.7%)** | 16 / 3820<br>(0.4%) | 280 /<br>2785<br>(10%) | **453 /**<br>**1935**<br>**(23%)** | **207 / 1558**<br>**(13%)** | **1,557 /**<br>**3116**<br>**(50%)** |  |  |

<sup>1</sup>n / N (%);
\*Pearson's Chi-squared test; p-values were calculated via Monte Carlo simulation (n=2000 replicates).
Bolded columns indicate classes with high diagnostic heterogeneity.
SCZ-Mood = schizophrenia with moderate mood swings; SAD-Bip = schizoaffective bipolar subtype;
SAD-Dep = schizoaffective depressed subtype; SCZ = schizophrenia

### Relationships to Demographic, Etiologic, and Clinical Correlates

All LCA constructs exhibited significant associations with demographic, etiologic, and clinical correlates (Tables S14-17). For example, Class 3 (SAD-Bip) had the earliest age of onset, the highest proportion of individuals with a family history of schizophrenia, and the highest endorsements of poor work and social adjustments. That said, the magnitude of inter-class differences for most of these correlates were modest and comparable to what was observed for established diagnosis (Figure 1). Neither the sex-specific or symptom-focused LCA solutions showed a substantial change in association strength compared to the best-fitting primary LCA solution that incorporated all participants and indicators. As expected, the predictive power of the composite phenotypes (i.e., established diagnosis and estimated latent class membership) outperformed individual symptom indicators (Figure 1).

**Figure 1.**
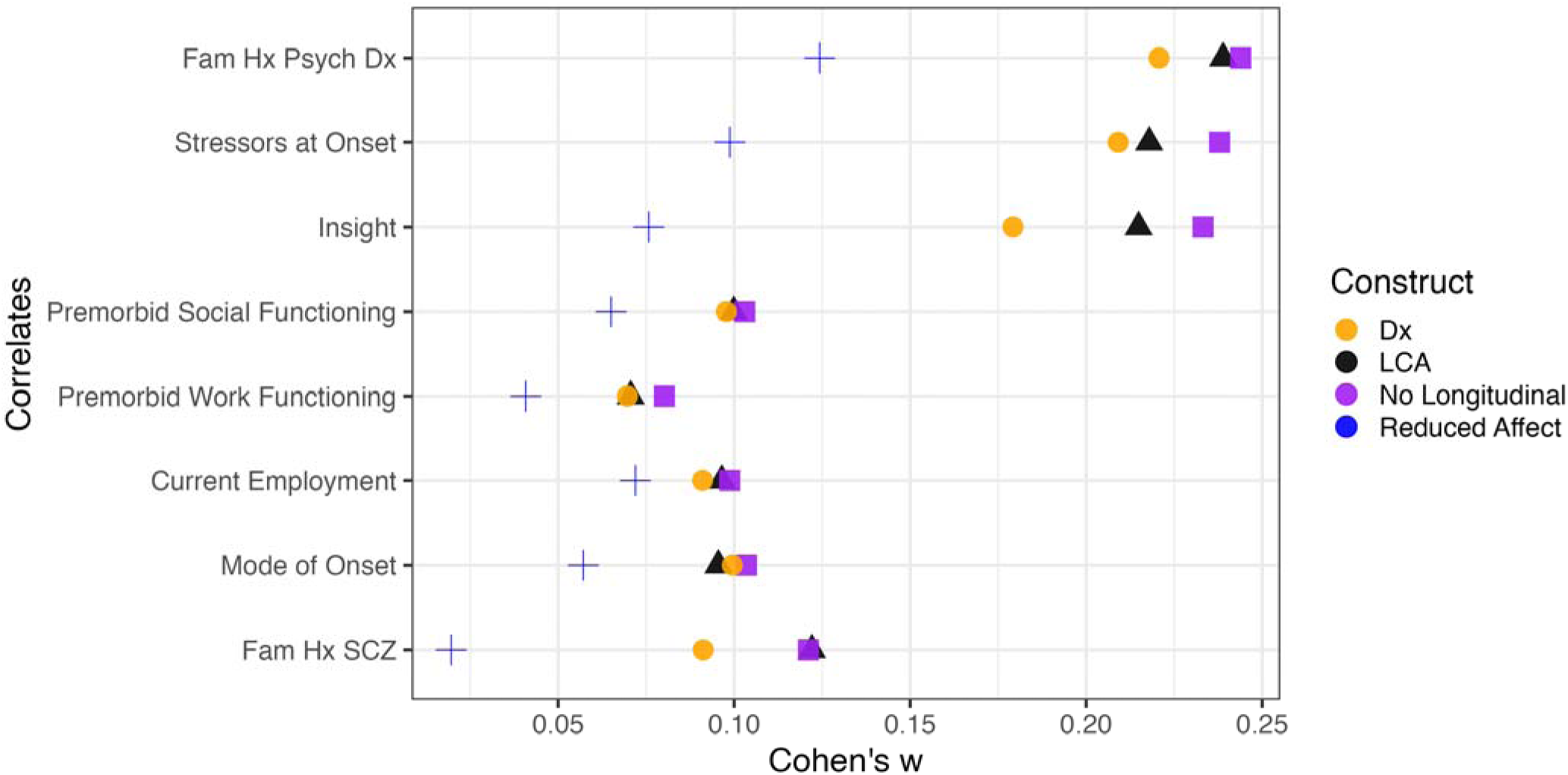
Comparison of Association Strength Between Estimated Latent Class Constructs and Established DSM Diagnosis and Clinical and Social Correlates. Effect sizes for the associations between established DSM diagnoses and latent class assignments for the primary (black triangle) and symptom-focused (i.e., no longitudinal course of illness items) analyses (purple square) are shown. There are no appreciable differences in predictive validity between either LCA construct and DSM diagnoses. To facilitate comparing these composite constructs, a single symptom indicator (reduced affect; blue +) is also included. As expected both the LCA constructs and DSM diagnoses, which encompass many symptoms, show stronger predictive validity with clinical and social functioning correlates than an individual symptom. Reduced affect (blue +) was chosen a priori given the strong association of negative symptoms with clinical outcomes. For completeness, several other single symptom indicators were also tested and they uniformed showed comparable magnitudes of association as reduced affect.

## Discussion

The primary objective of this study was to apply latent class analysis to a large, multi-diagnosis cohort of patients to derive transdiagnostic empirical phenotypes based on a symptom-level indicators. To our knowledge, this study represents the largest case-only latent class analysis of psychotic and affective symptoms to date. Subsequent analyses evaluated the robustness and external validity of the data-driven class assignments. Collectively, the six classes produced by the best-fitting LCA solution had distinct symptom profiles that captured combinations of mood lability and psychosis at varying severity levels and were generalizable across the sexes. Significant inter-class differences were observed for a small number of clinical characteristics and participant demographics. Specifically, the latent classes differed most substantially by illness course, the presence of impaired insight, and (in the full cohort analysis) the proportion of female participants. Subtle differences in clinical features and etiologic factors like age of onset, family history of schizophrenia, and premorbid work adjustment were present, but the effect sizes were modest. Latent class assignment for the primary and sex-specific analyses correlated with participants’ established diagnoses. Similar to other studies^8^, latent class membership was not significantly tied to disorder subtype.

The two secondary analyses provide important insight into the generalizability of the latent class symptom profiles and the applicability of LCA for future research endeavors. To our knowledge, no other transdiagnostic LCA study of psychosis has performed sex-stratified analyses. Our results suggest that LCA-derived class profiles based on OPCRIT symptom indicators may vary in prevalence by sex but not content. This observation is especially encouraging given that case-only cohorts of psychotic disorders are unlikely to be sex-balanced due to documented differences in diagnosis rates by sex (i.e., male excess for schizophrenia^29^).

The sensitivity analysis lacking course of illness indicators complements and extends prior work by Peralta and Cuesta (2007)^5^, which examined the impact of using lifetime indicator ratings compared to index episode ratings. In our study, we observed markedly increased within-class diagnostic heterogeneity combined with the decreased symptom profile distinctiveness, but these changes did not translate into significantly weaker associations with external validators, raising interesting questions about the value of the external indicators for differentiating patient clusters. Indeed, all of the LCA constructs showed comparable predictive validity to each other and to DSM diagnoses for clinical, etiological, and demographic factors. Overall, this analysis reaffirmed the importance of items that capture illness course and the relative timing and intensity of psychotic versus affective symptoms for generating latent classes that correlate with DSM diagnoses. Without their presence in the model, the relationship between latent class symptom profiles and established diagnoses deteriorated. This observation aligns with Kraepelin’s observations about the importance of illness course for distinguishing dementia praecox from manic depressive illness and underscores the importance for studies to collect this information and rely solely on instruments that only assess symptoms.

Most of the other large person-centered studies of psychotic illnesses have focused on a single disorder and so offer limited insight for contextualizing the results from this study^30–32^. When considering the other large, multi-diagnosis cohorts^10^ ^5^ ^8^ ^9^, there are three notable observations. First, latent class profiles based on OPCRIT items appear to generate replicable symptom profiles that generalize across the sexes. Three of six classes identified in this study resemble classes described by at least two previous studies (i.e., Class 6 (SCZ), Class 5 (SAD-Dep), and Class 3 (SAD-Bip)). Detailed comparisons of these profiles and their counterpart symptom profiles in other studies are provided in Supplemental File 1. Second, symptom-based clustering (whether using LCA or another grouping algorithm) seems to recapitulate DSM-based diagnoses fairly well, especially if proxies of illness course are included as indicators, but the resulting groups lack distinct profiles of social, clinical, or etiological factors. Third, the effect sizes for associations between DSM diagnoses and data-driven clusters with important clinical correlates were comparable to each other.

Care must be taken to consider the findings within the context of several limitations. One, LCA-based clustering is item-driven, comparative, and uneven in statistical confidence. Overall, we observed exceptionally high mean posterior probabilities (0.871 to 0.940) for all classes except SCZ-Mood (Class 1). Class 1’s low average posterior probability was driven by ∼1700 individuals (approximately half of those assigned to Class 1) who had a nontrivial amount of missing data and so would likely have had low posterior probabilities no matter the class solution. Two, no gold standard approach exists for determining the optimal number of classes to extract. Three, our item set had relatively sparse coverage of negative symptoms (i.e., only two indicators). This limitation is somewhat ameliorated by the fact that we used as close to the same OPCRIT items and coding strategy of prior studies^5,10^ as possible to promote cross-study comparability. Four, the phenotyping resolution for several of the clinical validators was suboptimal, which may have reduced our ability to detect differential class associations. For example, we were unable to use OPCRIT 89 *Responsiveness to Antipsychotic Medications* because both nonresponse and non-exposure were scored as zero. Five, the severe end of the phenotypic range for schizophrenia, schizoaffective disorder, and bipolar I disorder may be overrepresented in the GPC, undercutting its generalizability. Nearly all of the cases for all three disorders deteriorated from premorbid level of functioning, experienced auditory hallucinations, and reported significant impairment while ill (e.g., inpatient hospitalization). Additionally, the bipolar I cases with psychosis were intentionally oversampled, which could reduce the total number of unique symptom profiles observable in this cohort.

If the ultimate goal of using data-driven approaches is to refine the overlapping clinical presentations of psychotic disorders into informative subpopulations that reflect biological correlates, genetic liability, or functional measures (e.g., employment adjustment, cognitive functioning), more work is needed. Specifically, we must carefully evaluate the differential benefits of dimensional and categorical approaches compared to established diagnostic definitions, which generally are more readily accessible for large-scale cohorts via electronic health records compared to symptom-based phenotypes that require clinical interviews. These data-driven constructs must be thoroughly vetted for generalizability and comprehensively evaluated to determine if/how they relate to neurobiological biomarkers, aggregate measures of common and rare genetic variation, and clinical characteristics. Phenotype-only studies can infer a relationship with genetic factors based on associations with family history and clinical characteristics strongly associated with genetic liability (e.g., age of onset^33^), but explicit testing using single and multi-ancestry methods are needed.

## Supporting information

Coding environment

Description of replicated symptom profiles

## Data Availability

Individuals interested in data access to the Genomic Psychiatry Cohort should contact either the study PIs or seek authorized access through dbGaP (dbGaP Study Accession: phs001020.v2.p1). The analysis scripts are available on the Open Science Framework (scripts: https://osf.io/y6ftw, preregistration: https://osf.io/au2hg).

https://osf.io/au2hg

## Data Availability

https://osf.io/au2hg

## Data Availability

https://osf.io/au2hg

## Acknowledgements and Disclosures

The Genome Psychiatry Cohort sample ascertainment was supported by NIH grants MH085548 and MH085542 and the Stanley Center for Psychiatric Research at the Broad Institute. The whole genome sequencing of the GPC study was supported by NHGRI/NIH 5U54HG003067, a philanthropic gift to the Stanley Center for Psychiatric Research, and NIH grants NIMH/NIH R01 MH086873 and NIMH/NIH R01MH085548, as well as, NIMH/NIH U01MH105653 (Boehnke), NIMH/NIH U01MH105573 (Patos), and NIMH/NIH U01MH105641 (McCarroll). Additional support for the GPC comes from NIMH/NIH R01MH104964 (mPIs Pato, Pato, Fanous, Bigdeli) and NIMH/NIH R01MH123451 (mPIs Pato, Pato, Fanous, Bigdeli). DML was supported by NIMH/NIH K01MH131847.

## Data Availability Statement

Individuals interested in GPC data access should contact either the study PIs or seek authorized access through dbGaP (dbGaP Study Accession: phs001020.v2.p1). The analysis scripts are available on the Open Science Framework (scripts: https://osf.io/y6ftw, preregistration: https://osf.io/au2hg).

## Consortium Acknowledgements

### Genomic Psychiatry Cohort (GPC) Investigators

Michele T Pato MD ^1,2^, Carlos N Pato, MD, PhD ^1,2^, Tim B Bigdeli, PhD ^1,2,3^, Ayman H Fanous, MD ^1,2,3^, Steven A McCarroll, PhD ^4,5^, Peter F Buckley, MD ^6^, Mark J. Daly ^7,8,9,5^, James A Knowles MD, PhD ^2,10^, Douglas S Lehrer, MD ^11^, Dolores Malaspina, MD, MSPH ^12,13^, Mark H Rapaport, MD ^14^, Jeffrey J Rakofsky, MD ^14^, Janet L Sobell, PhD ^15^, Giulio Genovese, PhD ^4,5^, Penelope Georgakopoulos, DrPH ^2^, Jacquelyn L Meyers, PhD ^1^, Roseann E Peterson, PhD ^6^, Helena Medeiros, MSW ^2^, Jorge Valderrama, PhD ^1,2^, Eric D Achtyes, MD ^16^, Roman Kotov, PhD ^17^, Colony Abbott, MPH ^16^, Maria Helena Azevedo, PhD ^18^, Richard A Belliveau, Jr, BA ^4^, Elizabeth Bevilacqua, BS ^19^, Evelyn J Bromet, PhD ^17^, William Byerley, MD ^20^, Celia Barreto Carvalho, PhD ^21^, Sinéad B Chapman, MS ^4^, Lynn E DeLisi, MD ^22,23^, Ashley L Dumont, BASc ^4^, Colm O’Dushlaine, PhD ^4^, Laura J Fochtmann, MD ^17^, Diane Gage ^4^, James L Kennedy, MD ^24^, Becky Kinkead, PhD ^14^, Antonio Macedo, PhD ^18^, Jennifer L Moran, PhD ^4^, Christopher P Morley, PhD ^25-27^, Mantosh J Dewan, MD ^27^, James Nemesh ^4^, Diana O Perkins, MD, MPH ^28^, Shaun M Purcell, PhD ^4,29^, Edward M Scolnick, MD ^4^, Brooke M Sklar, MA ^15^, Pamela Sklar, MD, PhD ^12,13^, Jordan W Smoller, MD, ScD ^4,23,30,31^, Patrick F Sullivan, MD, FRANZCP ^28,32^, Humberto Nicolini, MD ^33^, Conrad O Iyegbe, PhD ^34^, Fabio Macciardi, MD, PhD ^35^, Stephen R Marder, MD ^36,37^, Michael A Escamilla, MD ^38^, Ruben C Gur, PhD ^39-41^, Raquel E Gur, MD, PhD ^39-41^, Tiffany A Greenwood, PhD ^42^, David L Braff, MD ^42,43^, Marquis P Vawter, PhD, MA, MS ^35^

^1^ Department of Psychiatry and Behavioral Sciences and ^2^ Institute for Genomic Health, SUNY Downstate Medical Center, Brooklyn, NY, USA; ^3^ VA New York Harbor Healthcare System, Brooklyn, NY, USA; ^4^ Stanley Center for Psychiatric Research, Broad Institute of MIT and Harvard, Cambridge, MA, USA; ^5^ Department of Genetics, Harvard Medical School, Boston, MA, USA; ^6^ School of Medicine, Virginia Commonwealth University, Richmond, VA, USA; ^7^ Institute for Molecular Medicine Finland (FIMM), University of Helsinki, Helsinki, Finland; ^8^ Analytic and Translational Genetics Unit, Massachusetts General Hospital, Boston, MA, USA; ^9^ Program in Medical and Population Genetics, Broad Institute of Harvard and MIT, Cambridge, MA, USA; ^10^ Department of Cell Biology, SUNY Downstate Medical Center, Brooklyn, NY, USA; ^11^ Department of Psychiatry, Wright State University, Dayton, OH, USA; ^12^ Departments of Psychiatry and ^13^ Genetics & Genomics, Icahn School of Medicine at Mount Sinai, NY, USA; ^14^ Department of Psychiatry and Behavioral Sciences, Emory University, Atlanta, GA, USA; ^15^ Department of Psychiatry & Behavioral Sciences, University of Southern California, Los Angeles, CA, USA; ^16^ Western Michigan University Homer Stryker M.D. School of Medicine, Kalamazoo, MI, USA; ^17^ Department of Psychiatry, Stony Brook University, Stony Brook, NY, USA; ^18^ Institute of Medical Psychology, Faculty of Medicine, University of Coimbra, Coimbra, PT; ^19^ Beacon Health Options, Boston, MA, USA; ^20^ Department of Psychiatry, University of California, San Francisco, CA, USA; ^21^ Faculty of Social and Human Sciences, University of Azores, PT; ^22^ VA Boston Healthcare System, Brockton, MA, USA; ^23^ Department of Psychiatry, Harvard Medical School, Boston, MA, USA; ^24^ Neurogenetics Laboratory, Campbell Family Mental Health Research Institute, Centre for Addiction and Mental Health; Department of Psychiatry, University of Toronto, ON, CA; ^25^ Departments of Public Health and Preventive Medicine, ^26^ Family Medicine, and ^27^ Psychiatry and Behavioral Sciences, State University of New York, Upstate Medical University, Syracuse, NY, USA; ^28^ Department of Psychiatry, University of North Carolina, Chapel Hill, NC, USA; ^29^ Department of Psychiatry, Brigham and Women’s Hospital, Boston, MA, USA; ^30^ Department of Psychiatry, Massachusetts General Hospital, Boston, MA, USA; ^31^ Department of Epidemiology, Harvard T.H. Chan School of Public Health, Boston, MA, USA; ^32^ Medical Epidemiology and Biostatistics, Karolinska Institutet, Solna, SE; ^33^ Carracci Medical Group, Mexico City, MX; ^34^ Department of Psychosis Studies, King’s College London, London, UK; ^35^ Department of Psychiatry and Human Behavior, University of California, Irvine, CA, USA; ^36^ Department of Psychiatry and Biobehavioral Sciences and ^37^ Semel Institute for Neuroscience and Human Behavior, Geffen School of Medicine, University of California Los Angeles, Los Angeles, CA, USA; ^38^ Department of Psychiatry, University of Texas Rio Grande Valley School of Medicine; ^39^ Departments of Psychiatry and ^40^ Child & Adolescent Psychiatry and ^41^ Lifespan Brain Institute, University of Pennsylvania Perelman School of Medicine and Children’s Hospital of Philadelphia, Philadelphia, PA, USA; ^42^ Department of Psychiatry, University of California, La Jolla, San Diego, CA, USA; ^43^ VISN-22 Mental Illness, Research, Education and Clinical Center (MIRECC), VA San Diego Healthcare System, San Diego, CA, USA

