## Supplemental Tables for "Latent class analysis of symptoms across schizophrenia, schizoaffective disorder, and bipolar I disorder"

### Table S1. OPCRIT Item Coding

| **Variable** | **OPCRIT Number and Title** | **Threshold** |
| --- | --- | --- |
| Positive thought disorder | 26 (Speech difficult to understand), 27 (Speech incoherent), or 28 (Positive formal thought disorder) | (0 vs 1/2) |
| Negative thought disorder | 29 (Negative formal thought disorder) | (0 vs 1/2) |
| Affective symptoms predominate | 52 (Relationship between psychotic and affective symptoms) | (0-2) vs 3 |
| Reduced affect | 33 (Blunted affect) or 32 (Restricted affect) | (0 vs 1/2) |
| Persecutory delusions | 54 (Persecutory delusions) | (0 vs 1/2) |
| Schneiderian delusions | 66 (Thought insertion), 67 (Thought withdrawal), or 68 (Thought broadcast) | (0 vs 1/2) |
| Schneiderian hallucinations | 73 (Third person) or 74 (Running commentary) | (0 vs 1/2) |
| Other auditory hallucinations | 75 (Command voices) or 77 (Hallucinations) | (0 vs 1/2) |
| Deterioration | 88 (Deterioration from premorbid level of functioning) | (0 vs 1) |
| Elevated mood | 35 (Elevated mood) | (0/1 vs 2/3) |
| Excessive activity | 19 (Excessive activity) | (0/1 vs 2/3) |
| Reckless activity | 20 (Reckless activity) | (0/1 vs 2/3) |
| Pressured speech | 30 (Pressured speech) | (0/1 vs 2/3) |
| Dysphoria | 37 (Dysphoria) | (0/1 vs 2/3) |
| Psychomotor change | 23 (Agitated activity) or 24 (Slowed) | (0/1 vs 2/3) |
| Energy | 25 (Loss of energy or fatigue) | (0/1 vs 2/3) |
| Excessive self-reproach | 42 (Excessive self-reproach) | (0/1 vs 2/3) |
| Reduction in weight or appetite | 48 (Decreased appetite) or 49 (Weight loss) | (0/1 vs 2/3) |
| Course | 90 (Course of disorder) | (1-3 vs 4/5) |
| Item coding based on *Kendler et al. (1998)* | | |

### Table S2. LCA Model Fit Indices for Primary Analysis

| **No. Classes** | **Parameters** | **DF** | **-2LL** | **AIC** | **BIC** | **X^2^** |
| --- | --- | --- | --- | --- | --- | --- |
| 2 | 39 | 17680 | -126794.07 | 253666.13 | 253969.64 | 5147571.26 |
| 3 | 59 | 17660 | -123490.45 | 247098.9 | 247558.07 | 3552405.09 |
| 4 | 79 | 17640 | -122892.45 | 245942.91 | 246557.72 | 3664957.2 |
| 5 | 99 | 17620 | -121784.25 | 243766.5 | 244536.96 | 4278821.81 |
| 6 | 119 | 17600 | -120747.52 | 241733.03 | 242659.14 | 2212754.22 |

Notes: DF = degrees of freedom; LL = log likelihood; AIC = Akaike Information Criterion; BIC = Bayesian Information Criterion; X^2^ = Chi Square

### Table S3. Item Endorsement by Class Membership for 6-Class Solution for Primary Analysis

|  | **Class 1: SCZ-Mood** | **Class 2: Psychotic Bipolar** | **Class 3: SAD-Bip** | **Class 4: Nonpsychotic Bipolar** | **Class 5: SAD-Dep** | **Class 6: SCZ** |
| --- | --- | --- | --- | --- | --- | --- |
| Proportion of Sample | 0.202 | 0.173 | 0.162 | 0.058 | 0.184 | 0.220 |
| Positive thought disorder | 0.386 | 0.246 | 0.454 | 0.055 | 0.332 | 0.493 |
| Negative thought disorder | 0.067 | 0.075 | 0.227 | 0.004 | **0.773** | **0.715** |
| Affective symptoms predominate | 0.017 | **0.993** | 0.073 | 0.027 | 0.000 | 0.000 |
| Reduced affect | 0.130 | 0.263 | 0.376 | 0.078 | **0.931** | **0.897** |
| Persecutory delusions | **0.808** | **0.629** | **0.894** | 0.000 | **0.887** | **0.883** |
| Schneiderian delusions | **0.579** | 0.340 | **0.757** | 0.000 | **0.744** | **0.678** |
| Schneiderian hallucinations | **0.535** | 0.304 | **0.772** | 0.000 | **0.707** | **0.655** |
| Other auditory hallucinations | **0.980** | **0.894** | **0.992** | 0.012 | **0.981** | **0.974** |
| Deterioration from premorbid condition | **0.989** | **0.941** | **1.000** | **0.889** | **1.000** | **1.000** |
| Elevated mood | 0.231 | **0.915** | **0.805** | **0.908** | 0.014 | 0.010 |
| Excessive activity | 0.466 | **0.877** | **0.939** | **0.898** | 0.093 | 0.029 |
| Reckless activity | 0.478 | **0.836** | **0.897** | **0.822** | 0.116 | 0.025 |
| Pressured speech | 0.469 | **0.902** | **0.928** | **0.920** | 0.049 | 0.017 |
| Dysphoria | 0.424 | **0.920** | **0.921** | **0.938** | **0.992** | 0.117 |
| Psychomotor change | **0.579** | **0.876** | **0.983** | **0.807** | **0.855** | 0.188 |
| Tired | **0.532** | **0.926** | **0.977** | **0.946** | **0.953** | 0.005 |
| Excessive self-reproach | 0.435 | **0.788** | **0.865** | **0.799** | **0.527** | 0.051 |
| Reduction in weight/appetite | 0.485 | **0.672** | **0.711** | **0.625** | 0.418 | 0.078 |
| Chronic course | **0.976** | 0.212 | **0.913** | 0.062 | **0.998** | **0.996** |
| *Endorsement frequencies >= 50% are bolded.*  SCZ-Mood = Schizophrenia with moderate mood swings; SAD-Bip = schizoaffective bipolar subtype; SAD-Dep = schizoaffective depressed subtype; SCZ = schizophrenia | | | | | | |

### Table S4. Relationship Between Established Diagnosis and OP52 Rating

| **Characteristic** | **Overall**  N = 16,014*^1^* | **0. No co-occurrence**.  N = 5021 (31%)*^1^* | **1. Psychotic symptoms dominate. Occasional affective disturbances may occur.**  N = 4972 (31%)*^1^* | **2. Psychotic and affective symptoms are balanced.**  N = 2706 (17%)*^1^* | **3. Affective symptoms dominate. Psychotic symptoms may also occur.**  N = 3315 (21%)*^1^* | **p-value*** | **Cohen’s w** |
| --- | --- | --- | --- | --- | --- | --- | --- |
| **Dx** |  |  |  |  |  | **<0.0005** | **1.3** |
| Schizophrenia | 8,856 / 16014 (55%) | 3,871 / 5021 (77%) | 4,964 / 4972 (100%) | 15 / 2706 (0.6%) | 6 / 3315 (0.2%) |  |  |
| Bipolar I | 4,462 / 16014 (28%) | 1,145 / 5021 (23%) | 3 / 4972 (<0.1%) | 7 / 2706 (0.3%) | 3,307 / 3315 (100%) |  |  |
| Schizoaffective | 2,696 / 16014 (17%) | 5 / 5021 (<0.1%) | 5 / 4972 (0.1%) | 2,684 / 2706 (99%) | 2 / 3315 (<0.1%) |  |  |
| *^1^*n / N (%); *Pearson's Chi-squared test; p-values were calculated via Monte Carlo simulation (n=2000 replicates). | | | | | |  |  |

### Table S5. Relationship Between Established Diagnosis and OP90 Rating

| **Characteristic** | **Overall** N = 15,988*^1^* | **1. Single episode with good recovery**  N = 52 (0.3%)*^1^* | **2. Multiple episodes with good recovery between**  N = 532 (3.3%)*^1^* | **3. Multiple episodes with partial recovery between**  N = 3117 (19%)*^1^* | **4. Gradual onset over period up to six months**  N = 7222 (45%)*^1^* | **5. Insidious onset over period greater than six months**  N = 5065 (32%)*^1^* | **p-value*** | **Cohen’s w** |
| --- | --- | --- | --- | --- | --- | --- | --- | --- |
| **Dx** |  |  |  |  |  |  | **<0.0005** | **0.79** |
| Schizophrenia | 8,846 / 15988 (55%) | 20 / 52 (38%) | 13 / 532 (2.4%) | 36 / 3117 (1.2%) | 4,823 / 7222 (67%) | 3,954 / 5065 (78%) |  |  |
| Bipolar I | 4,454 / 15988 (28%) | 31 / 52 (60%) | 509 / 532 (96%) | 2,850 / 3117 (91%) | 821 / 7222 (11%) | 243 / 5065 (4.8%) |  |  |
| Schizoaffective | 2,688 / 15988 (17%) | 1 / 52 (1.9%) | 10 / 532 (1.9%) | 231 / 3117 (7.4%) | 1,578 / 7222 (22%) | 868 / 5065 (17%) |  |  |
| *^1^*n / N (%); *Pearson’s Chi-squared test; p-values were calculated via Monte Carlo simulation (n=2000 replicates). | | | | | | |  |  |

### Table S6. LCA Model Fit Statistics for 2-7 Classes - Female Subset Only

| **No. Classes** | **Parameters** | **DF** | **-2LL** | **AIC** | **BIC** | **X^2^** |
| --- | --- | --- | --- | --- | --- | --- |
| 2 | 39 | 6286 | -51246.87 | 102571.74 | 102835.08 | 7251098.19 |
| 3 | 59 | 6266 | -49793.6 | 99705.2 | 100103.58 | 3007357.19 |
| 4 | 79 | 6246 | -48975.8 | 98109.61 | 98643.03 | 2545858.74 |
| 5 | 99 | 6226 | -48412.34 | 97022.68 | 97691.15 | 2416839.75 |
| 6 | 119 | 6206 | -48076.35 | 96390.69 | 97194.21 | 3299624.31 |
| 7 | 139 | 6186 | -47783.09 | 95844.18 | 96782.74 | 1447785.35 |

Notes: DF = degrees of freedom; LL = log likelihood; AIC = Akaike Information Criterion; BIC = Bayesian Information Criterion; X^2^ = Chi Square

### Table S7. LCA Model Fit Statistics for 2-6 Classes - Male Subset Only

| **No. Classes** | **Parameters** | **DF** | **-2LL** | **AIC** | **BIC** | **X^2^** |
| --- | --- | --- | --- | --- | --- | --- |
| 2 | 39 | 9685 | -74924.23 | 149926.46 | 150206.57 | 3093113.11 |
| 3 | 59 | 9665 | -73072.35 | 146262.7 | 146686.46 | 2671249.84 |
| 4 | 79 | 9645 | -71619 | 143396 | 143963.41 | 2612353.87 |
| 5 | 99 | 9625 | -70890.68 | 141979.37 | 142690.42 | 3010534.24 |
| 6 | 119 | 9605 | -70298.1 | 140834.2 | 141688.9 | 3004528.09 |

Notes: DF = degrees of freedom; LL = log likelihood; AIC = Akaike Information Criterion; BIC = Bayesian Information Criterion; X^2^ = Chi Square

### Table S8. Item Endorsement for Best Fitting LCA - Female Subset Only

|  | **Class 1: SCZ-Mood** | **Class 2: SAD-Dep** | **Class 3: Nonpsychotic Bipolar** | **Class 4: Bipolar** | **Class 5: SAD-Bip** |
| --- | --- | --- | --- | --- | --- |
| Proportion of Sample | 0.169 | 0.275 | 0.100 | 0.209 | 0.247 |
| Positive thought disorder | 0.445 | 0.370 | 0.065 | 0.214 | 0.400 |
| Negative thought disorder | 0.477 | **0.713** | 0.003 | 0.043 | 0.204 |
| Affective symptoms predominate | 0.002 | 0.001 | 0.054 | **0.998** | 0.242 |
| Reduced affect | **0.642** | **0.831** | 0.071 | 0.234 | 0.350 |
| Persecutory delusions | **0.843** | **0.894** | 0.017 | **0.561** | **0.874** |
| Schneiderian delusions | **0.559** | **0.748** | 0.000 | 0.257 | **0.708** |
| Schneiderian hallucinations | **0.550** | **0.724** | 0.000 | 0.207 | **0.732** |
| Other auditory hallucinations | **0.958** | **0.989** | 0.056 | **0.903** | **0.994** |
| Deterioration from premorbid condition | **0.997** | **1.000** | **0.880** | **0.923** | **0.996** |
| Elevated mood | 0.049 | 0.021 | **0.914** | **0.911** | **0.860** |
| Excessive activity | 0.162 | 0.082 | **0.901** | **0.875** | **0.910** |
| Reckless activity | 0.099 | 0.127 | **0.818** | **0.812** | **0.867** |
| Pressured speech | 0.154 | 0.077 | **0.925** | **0.896** | **0.935** |
| Dysphoria | 0.161 | **0.836** | **0.948** | **0.932** | **0.935** |
| Psychomotor change | 0.132 | **0.831** | **0.835** | **0.847** | **0.969** |
| Tired | 0.012 | **0.948** | **0.949** | **0.926** | **0.960** |
| Excessive self-reproach | 0.068 | **0.592** | **0.824** | **0.767** | **0.842** |
| Reduction in weight/appetite | 0.131 | 0.465 | **0.638** | **0.673** | **0.674** |
| Chronic course | **0.990** | **1.000** | 0.084 | 0.128 | **0.847** |
| *Endorsement frequencies >= 50% are bolded.*  SCZ-Mood = schizophrenia with moderate mood swings; SAD-Bip = schizoaffective bipolar subtype; SAD-Dep = schizoaffective depressed subtype | | | | | |

### Table S9. Item Endorsement for Best Fitting LCA - Male Subset Only

|  | **Class 1: SAD-Bip** | **Class 2: SCZ** | **Class 3: SCZ-Mood** | **Class 4: Psychotic Bipolar** | **Class 5: Nonpsychotic Bipolar** | **Class 6: SAD-Dep** |
| --- | --- | --- | --- | --- | --- | --- |
| Proportion of Sample | 0.175 | 0.294 | 0.165 | 0.156 | 0.046 | 0.164 |
| Positive thought disorder | **0.514** | 0.484 | 0.356 | 0.272 | 0.049 | 0.329 |
| Negative thought disorder | 0.271 | **0.707** | 0.017 | 0.082 | 0.004 | **0.979** |
| Affective symptoms predominate | 0.046 | 0.000 | 0.001 | **0.989** | 0.033 | 0.000 |
| Reduced affect | 0.423 | **0.883** | 0.263 | 0.283 | 0.089 | **0.992** |
| Persecutory delusions | **0.886** | **0.874** | **0.858** | **0.652** | 0.000 | **0.898** |
| Schneiderian delusions | **0.736** | **0.670** | **0.666** | 0.371 | 0.000 | **0.778** |
| Schneiderian hallucinations | **0.749** | **0.650** | **0.611** | 0.332 | 0.000 | **0.717** |
| Other auditory hallucinations | **0.989** | **0.977** | **0.987** | **0.887** | 0.021 | **0.977** |
| Deterioration from premorbid condition | **0.999** | **0.999** | **0.994** | **0.950** | **0.902** | **1.000** |
| Elevated mood | **0.857** | 0.023 | 0.033 | **0.909** | **0.884** | 0.014 |
| Excessive activity | **0.914** | 0.066 | 0.026 | **0.866** | **0.877** | 0.171 |
| Reckless activity | **0.873** | 0.070 | 0.101 | **0.850** | **0.817** | 0.169 |
| Pressured speech | **0.894** | 0.053 | 0.069 | **0.893** | **0.895** | 0.066 |
| Dysphoria | **0.840** | 0.118 | **0.597** | **0.893** | **0.918** | **0.961** |
| Psychomotor change | **0.947** | 0.120 | **0.755** | **0.877** | **0.765** | **0.875** |
| Tired | **0.891** | 0.000 | **0.858** | **0.907** | **0.927** | **0.936** |
| Excessive self-reproach | **0.780** | 0.066 | **0.709** | **0.782** | **0.764** | 0.404 |
| Reduction in weight/appetite | **0.667** | 0.088 | **0.666** | **0.670** | **0.601** | 0.317 |
| Chronic course | **0.916** | **0.994** | **0.990** | 0.250 | 0.062 | **0.998** |
| *Endorsement frequencies >= 50% are bolded.*  SCZ-Mood = schizophrenia with moderate mood swings; SAD-Bip = schizoaffective bipolar subtype; SAD-Dep = schizoaffective depressed subtype; SCZ = schizophrenia | | | | | | |

### Table S10. Item Endorsement for Sensitivity Analysis Without Illness Course Indicators

|  | **Class 1: Bipolar** | **Class 2: SCZ** | **Class 3: SCZ-Mood** | **Class 4: SAD-Dep** | **Class 5: Bipolar - Severe mania** | **Class 6: SAD-Bip** |
| --- | --- | --- | --- | --- | --- | --- |
| Proportion of Sample | 0.254 | 0.216 | 0.157 | 0.109 | 0.088 | 0.176 |
| Positive thought disorder | 0.135 | 0.482 | 0.358 | 0.258 | 0.494 | 0.438 |
| Negative thought disorder | 0.020 | **0.699** | **0.925** | 0.058 | 0.140 | 0.224 |
| Reduced affect | 0.156 | **0.882** | **0.985** | 0.292 | 0.213 | 0.419 |
| Persecutory delusions | 0.383 | **0.879** | **0.906** | **0.756** | **0.854** | **0.894** |
| Schneiderian delusions | 0.077 | **0.673** | **0.766** | **0.604** | **0.533** | **0.792** |
| Schneiderian hallucinations | 0.046 | **0.654** | **0.726** | **0.566** | **0.534** | **0.787** |
| Other auditory hallucinations | **0.637** | **0.974** | **0.984** | **0.987** | **0.966** | **1.000** |
| Deterioration from premorbid condition | **0.916** | **1.000** | **1.000** | **0.989** | **0.981** | **0.991** |
| Elevated mood | **0.948** | 0.009 | 0.013 | 0.216 | 0.437 | **0.876** |
| Excessive activity | **0.925** | 0.018 | 0.133 | 0.235 | **0.846** | **0.959** |
| Reckless activity | **0.861** | 0.018 | 0.133 | 0.305 | **0.762** | **0.920** |
| Pressured speech | **0.931** | 0.008 | 0.066 | 0.316 | **0.868** | **0.953** |
| Dysphoria | **0.956** | 0.127 | **0.951** | **0.943** | 0.104 | **0.996** |
| Psychomotor change | **0.854** | 0.068 | **0.880** | **0.744** | **0.708** | **0.980** |
| Tired | **0.955** | 0.013 | **0.953** | **0.875** | 0.057 | **0.978** |
| Excessive self-reproach | **0.801** | 0.077 | 0.463 | **0.715** | 0.066 | **0.870** |
| Reduction in weight/appetite | **0.657** | 0.106 | 0.348 | **0.675** | 0.154 | **0.714** |
| *Endorsement frequencies >= 50% are bolded.*  SCZ-Mood = schizophrenia with moderate mood swings; SAD-Bip = schizoaffective bipolar subtype; SAD-Dep = schizoaffective depressed subtype; SCZ = schizophrenia | | | | | | |

### Table S11. LCA Model Fit Statistics for Sensitivity Analysis Without Illness Course Indicators

| **No. Classes** | **Parameters** | **DF** | **-2LL** | **AIC** | **BIC** | **X^2^** |
| --- | --- | --- | --- | --- | --- | --- |
| 2 | 35 | 17684 | -117616.87 | 235303.74 | 235576.12 | 1209578.04 |
| 3 | 53 | 17666 | -115125.64 | 230357.28 | 230769.74 | 778299.21 |
| 4 | 71 | 17648 | -113189.11 | 226520.23 | 227072.78 | 514897.88 |
| 5 | 89 | 17630 | -112208.58 | 224595.15 | 225287.79 | 352675.4 |
| 6 | 107 | 17612 | -111331.83 | 222877.65 | 223710.37 | 361615.88 |
| 7 | 125 | 17594 | -111204.13 | 222658.26 | 223631.06 | 165673.58 |
| 8 | 143 | 17576 | -110882.77 | 222051.54 | 223164.42 | 238551.85 |

Notes: DF = degrees of freedom; LL = log likelihood; AIC = Akaike Information Criterion; BIC = Bayesian Information Criterion; X^2^ = Chi Square

### Table S12. Relationship Between Latent Class Assignment and Established Diagnosis - Female Only Sensitivity Analysis

| **Characteristic** | **Overall** N = 6,325*^1^* | **Class 1: SCZ-Mood**, N = 1071 (17%)*^1^* | **Class 2: SAD-Dep**, N = 1742 (28%)*^1^* | **Class 3: Nonpsychotic Bipolar**, N = 632 (10.0%)*^1^* | **Class 4: Bipolar**, N = 1319 (21%)*^1^* | **Class 5: SAD-Bip**, N = 1561 (25%)*^1^* | **p-value***^2^* | **Cohen’s w** |
| --- | --- | --- | --- | --- | --- | --- | --- | --- |
| **Dx** |  |  |  |  |  |  | **<0.0005** | **1.08** |
| Schizophrenia | 2,762 / 6325 (44%) | 1,051 / 1071 (98%) | 1,487 / 1742 (85%) | 7 / 632 (1.1%) | 5 / 1319 (0.4%) | 212 / 1561 (14%) |  |  |
| Bipolar I | 2,388 / 6325 (38%) | 4 / 1071 (0.4%) | 5 / 1742 (0.3%) | 616 / 632 (97%) | 1,309 / 1319 (99%) | 454 / 1561 (29%) |  |  |
| Schizoaffective | 1,175 / 6325 (19%) | 16 / 1071 (1.5%) | 250 / 1742 (14%) | 9 / 632 (1.4%) | 5 / 1319 (0.4%) | 895 / 1561 (57%) |  |  |
| *^1^*n / N (%) | | | | | | | |  |
| *^2^*Pearson's Chi-squared test; p-values were calculated via Monte Carlo simulation (n=2000 replicates).  SCZ-Mood = schizophrenia with moderate mood swings; SAD-Bip = schizoaffective bipolar subtype; SAD-Dep = schizoaffective depressed subtype | | | | | | | |  |

### Table S13. Relationship Between Latent Class Assignment and Established Diagnosis - Male Only Sensitivity Analysis

| **Characteristic** | **Overall** N = 9,724*^1^* | **Class 1: SAD-Bip**, N = 1702 (18%)*^1^* | **Class 2: SCZ**, N = 2863 (29%)*^1^* | **Class 3: SCZ-Mood**, N = 1603 (16%)*^1^* | **Class 4: Bipolar**, N = 1513 (16%)*^1^* | **Class 5: Nonpsychotic Bipolar**, N = 450 (4.6%)*^1^* | **Class 6: SAD-Dep**, N = 1593 (16%)*^1^* | **p-value***^2^* | **Cohen’s w** |
| --- | --- | --- | --- | --- | --- | --- | --- | --- | --- |
| **Dx** |  |  |  |  |  |  |  | **<0.0005** | **1.08** |
| Schizophrenia | 6,120 / 9724 (63%) | 416 / 1702 (24%) | 2,850 / 2863 (100%) | 1,399 / 1603 (87%) | 6 / 1513 (0.4%) | 2 / 450 (0.4%) | 1,447 / 1593 (91%) |  |  |
| Bipolar I | 2,078 / 9724 (21%) | 134 / 1702 (7.9%) | 1 / 2863 (<0.1%) | 4 / 1603 (0.2%) | 1,492 / 1513 (99%) | 447 / 450 (99%) | 0 / 1593 (0%) |  |  |
| Schizoaffective | 1,526 / 9724 (16%) | 1,152 / 1702 (68%) | 12 / 2863 (0.4%) | 200 / 1603 (12%) | 15 / 1513 (1.0%) | 1 / 450 (0.2%) | 146 / 1593 (9.2%) |  |  |
| *^1^*n / N (%) | | | | | | | | |  |
| *^2^*Pearson's Chi-squared test; p-values were calculated via Monte Carlo simulation (n=2000 replicates).  SCZ-Mood = schizophrenia with moderate mood swings; SAD-Bip = schizoaffective bipolar subtype; SAD-Dep = schizoaffective depressed subtype; SCZ = schizophrenia | | | | | | | | |  |

| Table S14. Clinical Correlates By Latent Class For LCA With All Indicators | | | | | | | | |
| --- | --- | --- | --- | --- | --- | --- | --- | --- |
| **Characteristic** | **Overall** N = 17,719*^1^* | **Class 1: SCZ-Mood**, N = 3584 (20%)*^1^* | **Class 2: Psychotic Bipolar**,  N = 3073 (17%)*^1^* | **Class 3: SAD-Bip**,  N = 2863 (16%)*^1^* | **Class 4: Nonpsychotic Bipolar**,  N = 1036 (5.8%)*^1^* | **Class 5: SAD-Dep**, N = 3262 (18%)*^1^* | **Class 6: SCZ**,  N = 3901 (22%)*^1^* | **p-value***** |
| **Clinical Presentation (OPCRIT 52)** |  |  |  |  |  |  |  | **<0.001** |
| 0. No co-occurrence | 5,021 / 16014 (31%) | 803 / 1903 (42%) | 1 / 3067 (<0.1%) | 129 / 2858 (4.5%) | 1,006 / 1035 (97%) | 100 / 3257 (3.1%) | 2,982 / 3894 (77%) |  |
| 1. Psychotic symptoms dominate. Occasional affective disturbances may occur. | 4,972 / 16014 (31%) | 764 / 1903 (40%) | 2 / 3067 (<0.1%) | 589 / 2858 (21%) | 1 / 1035 (<0.1%) | 2,719 / 3257 (83%) | 897 / 3894 (23%) |  |
| 2. Psychotic and affective symptoms are balanced. | 2,706 / 16014 (17%) | 303 / 1903 (16%) | 18 / 3067 (0.6%) | 1,932 / 2858 (68%) | 0 / 1035 (0%) | 438 / 3257 (13%) | 15 / 3894 (0.4%) |  |
| 3. Affective symptoms dominate. Psychotic symptoms may also occur. | 3,315 / 16014 (21%) | 33 / 1903 (1.7%) | 3,046 / 3067 (99%) | 208 / 2858 (7.3%) | 28 / 1035 (2.7%) | 0 / 3257 (0%) | 0 / 3894 (0%) |  |
| **Course of Illness**  **(OPCRIT 90)** |  |  |  |  |  |  |  |  |
| 1. Single episode with good recovery | 52 / 15988 (0.3%) | 11 / 1891 (0.6%) | 22 / 3065 (0.7%) | 1 / 2856 (<0.1%) | 11 / 1034 (1.1%) | 0 / 3252 (0%) | 7 / 3890 (0.2%) |  |
| 2. Multiple episodes with good recovery between | 532 / 15988 (3.3%) | 9 / 1891 (0.5%) | 311 / 3065 (10%) | 14 / 2856 (0.5%) | 195 / 1034 (19%) | 1 / 3252 (<0.1%) | 2 / 3890 (<0.1%) |  |
| 3. Multiple episodes with partial recovery between | 3,117 / 15988 (19%) | 25 / 1891 (1.3%) | 2,083 / 3065 (68%) | 233 / 2856 (8.2%) | 764 / 1034 (74%) | 5 / 3252 (0.2%) | 7 / 3890 (0.2%) |  |
| 4. Gradual onset over period up to six months | 7,222 / 15988 (45%) | 1,211 / 1891 (64%) | 501 / 3065 (16%) | 1,707 / 2856 (60%) | 44 / 1034 (4.3%) | 1,900 / 3252 (58%) | 1,859 / 3890 (48%) |  |
| 5. Insidious onset over period greater than six months | 5,065 / 15988 (32%) | 635 / 1891 (34%) | 148 / 3065 (4.8%) | 901 / 2856 (32%) | 20 / 1034 (1.9%) | 1,346 / 3252 (41%) | 2,015 / 3890 (52%) |  |
| **Age at onset (years)** | 20.5 (8.7) | 21.6 (9.0) | 19.5 (9.2) | 18.8 (8.6) | 19.6 (9.5) | 20.8 (8.2) | 21.9 (7.8) | **<0.001** |
| **Stressors at Onset** |  |  |  |  |  |  |  | **<0.001** |
| None present | 9,987 / 16010 (62%) | 1,275 / 1914 (67%) | 1,544 / 3069 (50%) | 1,517 / 2860 (53%) | 550 / 1036 (53%) | 2,068 / 3247 (64%) | 3,033 / 3884 (78%) |  |
| Present | 6,023 / 16010 (38%) | 639 / 1914 (33%) | 1,525 / 3069 (50%) | 1,343 / 2860 (47%) | 486 / 1036 (47%) | 1,179 / 3247 (36%) | 851 / 3884 (22%) |  |
| **Employment at onset** |  |  |  |  |  |  |  | **<0.001** |
| Employed at onset | 12,331 / 16004 (77%) | 1,421 / 1912 (74%) | 2,498 / 3064 (82%) | 2,253 / 2857 (79%) | 897 / 1033 (87%) | 2,405 / 3251 (74%) | 2,857 / 3887 (74%) |  |
| Not employed at onset | 3,673 / 16004 (23%) | 491 / 1912 (26%) | 566 / 3064 (18%) | 604 / 2857 (21%) | 136 / 1033 (13%) | 846 / 3251 (26%) | 1,030 / 3887 (26%) |  |
| **Premorbid work adjustment** |  |  |  |  |  |  |  | **<0.001** |
| Good premorbid work adjustment | 8,972 / 16002 (56%) | 1,100 / 1906 (58%) | 1,833 / 3064 (60%) | 1,474 / 2859 (52%) | 666 / 1035 (64%) | 1,803 / 3251 (55%) | 2,096 / 3887 (54%) |  |
| Poor premorbid work adjustment | 7,030 / 16002 (44%) | 806 / 1906 (42%) | 1,231 / 3064 (40%) | 1,385 / 2859 (48%) | 369 / 1035 (36%) | 1,448 / 3251 (45%) | 1,791 / 3887 (46%) |  |
| **Premorbid social adjustment** |  |  |  |  |  |  |  | **<0.001** |
| Good premorbid social adjustment | 8,482 / 16003 (53%) | 1,125 / 1911 (59%) | 1,729 / 3061 (56%) | 1,302 / 2858 (46%) | 653 / 1036 (63%) | 1,621 / 3251 (50%) | 2,052 / 3886 (53%) |  |
| Poor premorbid social adjustment | 7,521 / 16003 (47%) | 786 / 1911 (41%) | 1,332 / 3061 (44%) | 1,556 / 2858 (54%) | 383 / 1036 (37%) | 1,630 / 3251 (50%) | 1,834 / 3886 (47%) |  |
| **Insight** |  |  |  |  |  |  |  | **<0.001** |
| Insight impaired | 3,949 / 15993 (25%) | 640 / 1899 (34%) | 497 / 3068 (16%) | 540 / 2854 (19%) | 87 / 1034 (8.4%) | 735 / 3247 (23%) | 1,450 / 3891 (37%) |  |
| Insight present | 12,044 / 15993 (75%) | 1,259 / 1899 (66%) | 2,571 / 3068 (84%) | 2,314 / 2854 (81%) | 947 / 1034 (92%) | 2,512 / 3247 (77%) | 2,441 / 3891 (63%) |  |
| **Family Hx SCZ** |  |  |  |  |  |  |  | **<0.001** |
| Family history of schizophrenia | 4,798 / 15994 (30%) | 580 / 1907 (30%) | 888 / 3065 (29%) | 1,120 / 2855 (39%) | 215 / 1034 (21%) | 1,065 / 3251 (33%) | 930 / 3882 (24%) |  |
| No family history of schizophrenia | 11,196 / 15994 (70%) | 1,327 / 1907 (70%) | 2,177 / 3065 (71%) | 1,735 / 2855 (61%) | 819 / 1034 (79%) | 2,186 / 3251 (67%) | 2,952 / 3882 (76%) |  |
| **Family Hx Other** |  |  |  |  |  |  |  | **<0.001** |
| Family history of psychiatric disorder other than schizophrenia | 6,962 / 15981 (44%) | 771 / 1902 (41%) | 1,817 / 3069 (59%) | 1,478 / 2854 (52%) | 580 / 1035 (56%) | 1,266 / 3244 (39%) | 1,050 / 3877 (27%) |  |
| No family history | 9,019 / 15981 (56%) | 1,131 / 1902 (59%) | 1,252 / 3069 (41%) | 1,376 / 2854 (48%) | 455 / 1035 (44%) | 1,978 / 3244 (61%) | 2,827 / 3877 (73%) |  |
| *^1^*n / N (%); Mean (SD) | | | | | | | | |
| ***Pearson's Chi-squared test; Kruskal-Wallis rank sum test.  SCZ-Mood = schizophrenia with moderate mood swings; SAD-Bip = schizoaffective bipolar subtype; SAD-Dep = schizoaffective depressed subtype; SCZ = schizophrenia | | | | | | | | |

| Table S15. Clinical Correlates By Latent Class - Female Only | | | | | | | |
| --- | --- | --- | --- | --- | --- | --- | --- |
| **Characteristic** | **Overall** N = 5,251*^1^* | **Class 1: SCZ-Mood**, N = 1061 (20%)*^1^* | **Class 2: SAD-Dep**, N = 1333 (25%)*^1^* | **Class 3: Nonpsychotic Bipolar**, N = 634 (12%)*^1^* | **Class 4: Bipolar**, N = 625 (12%)*^1^* | **Class 5: SAD-Bip**, N = 1598 (30%)*^1^* | **p-value***^2^* |
| **Age at onset (years)** | 20.2 (9.2) | 21.4 (8.7) | 19.2 (9.0) | 19.6 (9.5) | 22.5 (9.7) | 19.4 (9.3) | **<0.001** |
| **Family Hx SCZ** |  |  |  |  |  |  | **<0.001** |
| Family history of schizophrenia | 1,802 / 5235 (34%) | 349 / 1057 (33%) | 554 / 1330 (42%) | 141 / 633 (22%) | 240 / 622 (39%) | 518 / 1593 (33%) |  |
| No family history of schizophrenia | 3,433 / 5235 (66%) | 708 / 1057 (67%) | 776 / 1330 (58%) | 492 / 633 (78%) | 382 / 622 (61%) | 1,075 / 1593 (67%) |  |
| **Family Hx Other** |  |  |  |  |  |  | **<0.001** |
| Family history of psychiatric disorder other than schizophrenia | 2,894 / 5237 (55%) | 452 / 1057 (43%) | 751 / 1327 (57%) | 372 / 634 (59%) | 294 / 622 (47%) | 1,025 / 1597 (64%) |  |
| No family history | 2,343 / 5237 (45%) | 605 / 1057 (57%) | 576 / 1327 (43%) | 262 / 634 (41%) | 328 / 622 (53%) | 572 / 1597 (36%) |  |
| **Course of Illness** |  |  |  |  |  |  |  |
| 1. Single episode with good recovery | 19 / 5233 (0.4%) | 0 / 1058 (0%) | 1 / 1330 (<0.1%) | 8 / 630 (1.3%) | 0 / 622 (0%) | 10 / 1593 (0.6%) |  |
| 2. Multiple episodes with good recovery between | 311 / 5233 (5.9%) | 1 / 1058 (<0.1%) | 5 / 1330 (0.4%) | 120 / 630 (19%) | 0 / 622 (0%) | 185 / 1593 (12%) |  |
| 3. Multiple episodes with partial recovery between | 1,630 / 5233 (31%) | 1 / 1058 (<0.1%) | 97 / 1330 (7.3%) | 446 / 630 (71%) | 0 / 622 (0%) | 1,086 / 1593 (68%) |  |
| 4. Gradual onset over period up to six months | 2,120 / 5233 (41%) | 623 / 1058 (59%) | 818 / 1330 (62%) | 42 / 630 (6.7%) | 396 / 622 (64%) | 241 / 1593 (15%) |  |
| 5. Insidious onset over period greater than six months | 1,153 / 5233 (22%) | 433 / 1058 (41%) | 409 / 1330 (31%) | 14 / 630 (2.2%) | 226 / 622 (36%) | 71 / 1593 (4.5%) |  |
| **Insight** |  |  |  |  |  |  | **<0.001** |
| Insight impaired | 963 / 5237 (18%) | 216 / 1057 (20%) | 262 / 1329 (20%) | 53 / 633 (8.4%) | 195 / 622 (31%) | 237 / 1596 (15%) |  |
| Insight present | 4,274 / 5237 (82%) | 841 / 1057 (80%) | 1,067 / 1329 (80%) | 580 / 633 (92%) | 427 / 622 (69%) | 1,359 / 1596 (85%) |  |
| **Employment at onset** |  |  |  |  |  |  | **<0.001** |
| Employed at onset | 4,210 / 5241 (80%) | 805 / 1058 (76%) | 1,064 / 1329 (80%) | 557 / 634 (88%) | 458 / 624 (73%) | 1,326 / 1596 (83%) |  |
| Not employed at onset | 1,031 / 5241 (20%) | 253 / 1058 (24%) | 265 / 1329 (20%) | 77 / 634 (12%) | 166 / 624 (27%) | 270 / 1596 (17%) |  |
| **Premorbid work adjustment** |  |  |  |  |  |  | **<0.001** |
| Good premorbid work adjustment | 3,139 / 5240 (60%) | 628 / 1059 (59%) | 713 / 1330 (54%) | 418 / 634 (66%) | 364 / 623 (58%) | 1,016 / 1594 (64%) |  |
| Poor premorbid work adjustment | 2,101 / 5240 (40%) | 431 / 1059 (41%) | 617 / 1330 (46%) | 216 / 634 (34%) | 259 / 623 (42%) | 578 / 1594 (36%) |  |
| **Premorbid social adjustment** |  |  |  |  |  |  | **<0.001** |
| Good premorbid social adjustment | 2,790 / 5240 (53%) | 520 / 1057 (49%) | 607 / 1331 (46%) | 400 / 634 (63%) | 340 / 624 (54%) | 923 / 1594 (58%) |  |
| Poor premorbid social adjustment | 2,450 / 5240 (47%) | 537 / 1057 (51%) | 724 / 1331 (54%) | 234 / 634 (37%) | 284 / 624 (46%) | 671 / 1594 (42%) |  |
| *^1^*Mean (SD); n / N (%) | | | | | | | |
| *^2^*Kruskal-Wallis rank sum test; Pearson's Chi-squared test  SCZ-Mood = schizophrenia with moderate mood swings; SAD-Bip = schizoaffective bipolar subtype; SAD-Dep = schizoaffective depressed subtype | | | | | | | |

| Table S16. Clinical Correlates By Latent Class - Male Only | | | | | | | | |
| --- | --- | --- | --- | --- | --- | --- | --- | --- |
| **Characteristic** | **Overall**  N = 9,724*^1^* | **Class 1: SAD-Bip**  N = 1702 (18%)*^1^* | **Class 2: SCZ**  N = 2863 (29%)*^1^* | **Class 3: SCZ-Mood**  N = 1603 (16%)*^1^* | **Class 4: Bipolar**  N = 1513 (16%)*^1^* | **Class 5: Nonpsychotic Bipolar**  N = 450 (4.6%)*^1^* | **Class 6: SAD-Dep**  N = 1593 (16%)*^1^* | **p-value***^2^* |
| **Age at onset (years)** | 20.3 (8.2) | 18.5 (8.0) | 21.3 (7.4) | 21.1 (8.5) | 19.5 (9.0) | 19.8 (9.7) | 20.6 (7.9) | **<0.001** |
| **Family Hx SCZ** |  |  |  |  |  |  |  | **<0.001** |
| Family history of schizophrenia | 2,723 / 9684 (28%) | 607 / 1696 (36%) | 675 / 2848 (24%) | 476 / 1595 (30%) | 389 / 1510 (26%) | 85 / 448 (19%) | 491 / 1587 (31%) |  |
| No family history of schizophrenia | 6,961 / 9684 (72%) | 1,089 / 1696 (64%) | 2,173 / 2848 (76%) | 1,119 / 1595 (70%) | 1,121 / 1510 (74%) | 363 / 448 (81%) | 1,096 / 1587 (69%) |  |
| **Family Hx Other** |  |  |  |  |  |  |  | **<0.001** |
| Family history of psychiatric disorder other than schizophrenia | 3,737 / 9672 (39%) | 819 / 1695 (48%) | 749 / 2843 (26%) | 599 / 1594 (38%) | 800 / 1511 (53%) | 237 / 449 (53%) | 533 / 1580 (34%) |  |
| No family history | 5,935 / 9672 (61%) | 876 / 1695 (52%) | 2,094 / 2843 (74%) | 995 / 1594 (62%) | 711 / 1511 (47%) | 212 / 449 (47%) | 1,047 / 1580 (66%) |  |
| **Course of Illness** |  |  |  |  |  |  |  |  |
| 1. Single episode with good recovery | 31 / 9679 (0.3%) | 3 / 1690 (0.2%) | 7 / 2852 (0.2%) | 6 / 1592 (0.4%) | 12 / 1509 (0.8%) | 3 / 449 (0.7%) | 0 / 1587 (0%) |  |
| 2. Multiple episodes with good recovery between | 218 / 9679 (2.3%) | 9 / 1690 (0.5%) | 3 / 2852 (0.1%) | 2 / 1592 (0.1%) | 121 / 1509 (8.0%) | 83 / 449 (18%) | 0 / 1587 (0%) |  |
| 3. Multiple episodes with partial recovery between | 1,481 / 9679 (15%) | 130 / 1690 (7.7%) | 6 / 2852 (0.2%) | 8 / 1592 (0.5%) | 999 / 1509 (66%) | 335 / 449 (75%) | 3 / 1587 (0.2%) |  |
| 4. Gradual onset over period up to six months | 4,589 / 9679 (47%) | 968 / 1690 (57%) | 1,396 / 2852 (49%) | 1,063 / 1592 (67%) | 293 / 1509 (19%) | 21 / 449 (4.7%) | 848 / 1587 (53%) |  |
| 5. Insidious onset over period greater than six months | 3,360 / 9679 (35%) | 580 / 1690 (34%) | 1,440 / 2852 (50%) | 513 / 1592 (32%) | 84 / 1509 (5.6%) | 7 / 449 (1.6%) | 736 / 1587 (46%) |  |
| **Insight** |  |  |  |  |  |  |  | **<0.001** |
| Insight impaired | 2,576 / 9679 (27%) | 365 / 1690 (22%) | 1,034 / 2853 (36%) | 457 / 1595 (29%) | 271 / 1509 (18%) | 44 / 448 (9.8%) | 405 / 1584 (26%) |  |
| Insight present | 7,103 / 9679 (73%) | 1,325 / 1690 (78%) | 1,819 / 2853 (64%) | 1,138 / 1595 (71%) | 1,238 / 1509 (82%) | 404 / 448 (90%) | 1,179 / 1584 (74%) |  |
| **Employment at onset** |  |  |  |  |  |  |  | **<0.001** |
| Employed at onset | 7,330 / 9691 (76%) | 1,324 / 1700 (78%) | 2,099 / 2853 (74%) | 1,171 / 1598 (73%) | 1,205 / 1506 (80%) | 379 / 447 (85%) | 1,152 / 1587 (73%) |  |
| Not employed at onset | 2,361 / 9691 (24%) | 376 / 1700 (22%) | 754 / 2853 (26%) | 427 / 1598 (27%) | 301 / 1506 (20%) | 68 / 447 (15%) | 435 / 1587 (27%) |  |
| **Premorbid work adjustment** |  |  |  |  |  |  |  | **<0.001** |
| Good premorbid work adjustment | 5,195 / 9686 (54%) | 846 / 1698 (50%) | 1,510 / 2849 (53%) | 872 / 1597 (55%) | 836 / 1508 (55%) | 287 / 449 (64%) | 844 / 1585 (53%) |  |
| Poor premorbid work adjustment | 4,491 / 9686 (46%) | 852 / 1698 (50%) | 1,339 / 2849 (47%) | 725 / 1597 (45%) | 672 / 1508 (45%) | 162 / 449 (36%) | 741 / 1585 (47%) |  |
| **Premorbid social adjustment** |  |  |  |  |  |  |  | **<0.001** |
| Good premorbid social adjustment | 5,098 / 9688 (53%) | 812 / 1698 (48%) | 1,512 / 2850 (53%) | 885 / 1596 (55%) | 824 / 1505 (55%) | 288 / 450 (64%) | 777 / 1589 (49%) |  |
| Poor premorbid social adjustment | 4,590 / 9688 (47%) | 886 / 1698 (52%) | 1,338 / 2850 (47%) | 711 / 1596 (45%) | 681 / 1505 (45%) | 162 / 450 (36%) | 812 / 1589 (51%) |  |
| *^1^*Mean (SD); n / N (%) | | | | | | | | |
| *^2^*Kruskal-Wallis rank sum test; Pearson's Chi-squared test  SCZ-Mood = schizophrenia with moderate mood swings; SAD-Bip = schizoaffective bipolar subtype; SAD-Dep = schizoaffective depressed subtype | | | | | | | | |

| Table S17. Clinical Correlates for Latent Classes Without Illness Course Indicators | | | | | | | | |
| --- | --- | --- | --- | --- | --- | --- | --- | --- |
| **Characteristic** | **Overall** N = 17,719*^1^* | **Class 1: Bipolar**  N = 4505 (25%)*^1^* | **Class 2: SCZ** N = 3820 (22%)*^1^* | **Class 3: SCZ-Mood**  N = 2785 (16%)*^1^* | **Class 4: SAD-Dep**  N = 1935 (11%)*^1^* | **Class 5: Bipolar-Severe Mania**  N = 1558 (8.8%)*^1^* | **Class 6: SAD-Bip**  N = 3116 (18%)*^1^* | **p-value***^2^* |
| **Age at onset (years)** | 20.5 (8.7) | 19.6 (9.3) | 22.0 (7.9) | 20.8 (8.1) | 20.4 (9.3) | 21.7 (8.8) | 18.5 (8.4) | **<0.001** |
| **Stressors at Onset** |  |  |  |  |  |  |  | **<0.001** |
| None present | 9,987 / 16010 (62%) | 1,478 / 2838 (52%) | 2,958 / 3804 (78%) | 1,834 / 2773 (66%) | 1,029 / 1930 (53%) | 1,150 / 1555 (74%) | 1,538 / 3110 (49%) |  |
| Present | 6,023 / 16010 (38%) | 1,360 / 2838 (48%) | 846 / 3804 (22%) | 939 / 2773 (34%) | 901 / 1930 (47%) | 405 / 1555 (26%) | 1,572 / 3110 (51%) |  |
| **Employment at onset** |  |  |  |  |  |  |  | **<0.001** |
| Employed at onset | 12,331 / 16004 (77%) | 2,407 / 2834 (85%) | 2,807 / 3806 (74%) | 2,048 / 2775 (74%) | 1,482 / 1932 (77%) | 1,140 / 1551 (74%) | 2,447 / 3106 (79%) |  |
| Not employed at onset | 3,673 / 16004 (23%) | 427 / 2834 (15%) | 999 / 3806 (26%) | 727 / 2775 (26%) | 450 / 1932 (23%) | 411 / 1551 (26%) | 659 / 3106 (21%) |  |
| **Premorbid work adjustment** |  |  |  |  |  |  |  | **<0.001** |
| Good premorbid work adjustment | 8,972 / 16002 (56%) | 1,785 / 2835 (63%) | 2,074 / 3806 (54%) | 1,526 / 2775 (55%) | 1,115 / 1930 (58%) | 899 / 1546 (58%) | 1,573 / 3110 (51%) |  |
| Poor premorbid work adjustment | 7,030 / 16002 (44%) | 1,050 / 2835 (37%) | 1,732 / 3806 (46%) | 1,249 / 2775 (45%) | 815 / 1930 (42%) | 647 / 1546 (42%) | 1,537 / 3110 (49%) |  |
| **Premorbid social adjustment** |  |  |  |  |  |  |  | **<0.001** |
| Good premorbid social adjustment | 8,482 / 16003 (53%) | 1,701 / 2833 (60%) | 2,028 / 3804 (53%) | 1,375 / 2776 (50%) | 1,059 / 1931 (55%) | 913 / 1550 (59%) | 1,406 / 3109 (45%) |  |
| Poor premorbid social adjustment | 7,521 / 16003 (47%) | 1,132 / 2833 (40%) | 1,776 / 3804 (47%) | 1,401 / 2776 (50%) | 872 / 1931 (45%) | 637 / 1550 (41%) | 1,703 / 3109 (55%) |  |
| **Insight** |  |  |  |  |  |  |  | **<0.001** |
| Insight impaired | 3,949 / 15993 (25%) | 369 / 2832 (13%) | 1,368 / 3806 (36%) | 684 / 2775 (25%) | 402 / 1921 (21%) | 648 / 1551 (42%) | 478 / 3108 (15%) |  |
| Insight present | 12,044 / 15993 (75%) | 2,463 / 2832 (87%) | 2,438 / 3806 (64%) | 2,091 / 2775 (75%) | 1,519 / 1921 (79%) | 903 / 1551 (58%) | 2,630 / 3108 (85%) |  |
| **Family Hx SCZ** |  |  |  |  |  |  |  | **<0.001** |
| Family history of schizophrenia | 4,798 / 15994 (30%) | 699 / 2831 (25%) | 922 / 3801 (24%) | 892 / 2777 (32%) | 675 / 1924 (35%) | 421 / 1554 (27%) | 1,189 / 3107 (38%) |  |
| No family history of schizophrenia | 11,196 / 15994 (70%) | 2,132 / 2831 (75%) | 2,879 / 3801 (76%) | 1,885 / 2777 (68%) | 1,249 / 1924 (65%) | 1,133 / 1554 (73%) | 1,918 / 3107 (62%) |  |
| **Family Hx Other** |  |  |  |  |  |  |  | **<0.001** |
| Family history of psychiatric disorder other than schizophrenia | 6,962 / 15981 (44%) | 1,655 / 2832 (58%) | 1,034 / 3794 (27%) | 1,035 / 2771 (37%) | 976 / 1926 (51%) | 554 / 1549 (36%) | 1,708 / 3109 (55%) |  |
| No family history | 9,019 / 15981 (56%) | 1,177 / 2832 (42%) | 2,760 / 3794 (73%) | 1,736 / 2771 (63%) | 950 / 1926 (49%) | 995 / 1549 (64%) | 1,401 / 3109 (45%) |  |
| **Clinical Presentation (OPCRIT52)** |  |  |  |  |  |  |  | **<0.001** |
| 0. No co-occurrence | 5,021 / 16014 (31%) | 1,029 / 2837 (36%) | 2,899 / 3811 (76%) | 174 / 2781 (6.3%) | 145 / 1923 (7.5%) | 755 / 1552 (49%) | 19 / 3110 (0.6%) |  |
| 1. Psychotic symptoms dominate. Occasional affective disturbances may occur. | 4,972 / 16014 (31%) | 50 / 2837 (1.8%) | 894 / 3811 (23%) | 2,311 / 2781 (83%) | 1,025 / 1923 (53%) | 333 / 1552 (21%) | 359 / 3110 (12%) |  |
| 2. Psychotic and affective symptoms are balanced. | 2,706 / 16014 (17%) | 193 / 2837 (6.8%) | 18 / 3811 (0.5%) | 281 / 2781 (10%) | 448 / 1923 (23%) | 210 / 1552 (14%) | 1,556 / 3110 (50%) |  |
| 3. Affective symptoms dominate. Psychotic symptoms may also occur. | 3,315 / 16014 (21%) | 1,565 / 2837 (55%) | 0 / 3811 (0%) | 15 / 2781 (0.5%) | 305 / 1923 (16%) | 254 / 1552 (16%) | 1,176 / 3110 (38%) |  |
| **Course of Illness (OPCRIT90)** |  |  |  |  |  |  |  | **<0.001** |
| 1. Single episode with good recovery | 52 / 15988 (0.3%) | 22 / 2831 (0.8%) | 10 / 3806 (0.3%) | 1 / 2779 (<0.1%) | 7 / 1919 (0.4%) | 11 / 1545 (0.7%) | 1 / 3108 (<0.1%) |  |
| 2. Multiple episodes with good recovery between | 532 / 15988 (3.3%) | 376 / 2831 (13%) | 4 / 3806 (0.1%) | 2 / 2779 (<0.1%) | 36 / 1919 (1.9%) | 36 / 1545 (2.3%) | 78 / 3108 (2.5%) |  |
| 3. Multiple episodes with partial recovery between | 3,117 / 15988 (19%) | 1,709 / 2831 (60%) | 12 / 3806 (0.3%) | 11 / 2779 (0.4%) | 254 / 1919 (13%) | 182 / 1545 (12%) | 949 / 3108 (31%) |  |
| 4. Gradual onset over period up to six months | 7,222 / 15988 (45%) | 549 / 2831 (19%) | 1,853 / 3806 (49%) | 1,520 / 2779 (55%) | 1,224 / 1919 (64%) | 752 / 1545 (49%) | 1,324 / 3108 (43%) |  |
| 5. Insidious onset over period greater than six months | 5,065 / 15988 (32%) | 175 / 2831 (6.2%) | 1,927 / 3806 (51%) | 1,245 / 2779 (45%) | 398 / 1919 (21%) | 564 / 1545 (37%) | 756 / 3108 (24%) |  |
| *^1^*Mean (SD); n / N (%) | | | | | | | | |
| *^2^*Kruskal-Wallis rank sum test; Pearson's Chi-squared test  SCZ-Mood = schizophrenia with moderate mood swings; SAD-Bip = schizoaffective bipolar subtype; SAD-Dep = schizoaffective depressed subtype; SCZ = schizophrenia | | | | | | | | |
