## Supplementary material for "Latent class analysis of symptoms across schizophrenia, schizoaffective disorder, and bipolar I disorder": Coding environment

**Supplemental File – Coding environment**

Analyses were conducted using the R Statistical language (version 4.2.1; R Core Team, 2022) on macOS Big Sur … 10.16, using the packages gt (version 0.9.0; Iannone R et al., 2023), report (version 0.5.7; Makowski D et al., 2023), rcompanion (version 2.4.30; Mangiafico SS, 2023), gtsummary (version 1.7.0; Sjoberg D et al., 2021), ggplot2 (version 3.4.2; Wickham H, 2016), dplyr (version 1.1.1; Wickham H et al., 2023), scales (version 1.2.1; Wickham H, Seidel D, 2022) and kableExtra (version 1.3.4; Zhu H, 2021).

### **Software References**

•	Iannone R, Cheng J, Schloerke B, Hughes E, Lauer A, Seo J (2023). *gt: Easily Create Presentation-Ready Display Tables*. R package version 0.9.0, [https://CRAN.R-project.org/package=gt](https://cran.r-project.org/package=gt).

•	Makowski D, Lüdecke D, Patil I, Thériault R, Ben-Shachar M, Wiernik B (2023). “Automated Results Reporting as a Practical Tool to Improve Reproducibility and Methodological Best Practices Adoption.” *CRAN*. <https://easystats.github.io/report/>.

•	Mangiafico SS (2023). *rcompanion: Functions to Support Extension Education Program Evaluation*. Rutgers Cooperative Extension, New Brunswick, New Jersey. version 2.4.30, [https://CRAN.R-project.org/package=rcompanion/](https://cran.r-project.org/package=rcompanion/).

•	R Core Team (2022). *R: A Language and Environment for Statistical Computing*. R Foundation for Statistical Computing, Vienna, Austria. [https://www.R-project.org/](https://www.r-project.org/).

•	Sjoberg D, Whiting K, Curry M, Lavery J, Larmarange J (2021). “Reproducible Summary Tables with the gtsummary Package.” *The R Journal*, *13*, 570-580. doi:10.32614/RJ-2021-053 <https://doi.org/10.32614/RJ-2021-053>, <https://doi.org/10.32614/RJ-2021-053>.

•	Wickham H (2016). *ggplot2: Elegant Graphics for Data Analysis*. Springer-Verlag New York. ISBN 978-3-319-24277-4, <https://ggplot2.tidyverse.org>.

•	Wickham H, François R, Henry L, Müller K, Vaughan D (2023). *dplyr: A Grammar of Data Manipulation*. R package version 1.1.1, [https://CRAN.R-project.org/package=dplyr](https://cran.r-project.org/package=dplyr).

•	Wickham H, Seidel D (2022). *scales: Scale Functions for Visualization*. R package version 1.2.1, [https://CRAN.R-project.org/package=scales](https://cran.r-project.org/package=scales).

•	Zhu H (2021). *kableExtra: Construct Complex Table with ‘kable’ and Pipe Syntax*. R package version 1.3.4, [https://CRAN.R-project.org/package=kableExtra](https://cran.r-project.org/package=kableExtra).
