## Supplementary material for "Latent class analysis of symptoms across schizophrenia, schizoaffective disorder, and bipolar I disorder": Description of replicated symptom profiles

**Supplemental File 1 – Detailed descriptions of the replicated symptom profiles**

Three symptom profiles identified in this study replicated prior findings. We shall review each in turn. First, all five studies reported extracting a class with a symptom profile that resembles Kraepelin’s descriptions of schizophrenia/dementia praecox. Alternatively named Class 6 - SCZ (schizophrenia; this study), “classic schizophrenia” (Kendler 1998), “schizophrenia” (Peralta and Cuesta 2003), “Cluster 3” (Boks 2007), and “Kraepelinian schizophrenia” (Derks 2012), the symptom profiles include high levels of psychotic symptoms, a chronic, deteriorating course of illness, low-to-absent affective symptoms, and poor outcome. This study observed that most of the individuals in this class reported no psychotic or affective illness in close relatives, which concurs with findings from Kendler et al. (1998).

Second, all five studies describe a class marked by high levels of a wide range of psychotic, manic, and depressive symptoms variously described as “schizobipolar” (Peralta and Cuesta), “bipolar-schizomania” (Kendler 1998), “affective psychosis” (Derks 2012), “Cluster 1” (Boks 2007), and “Class 3 - SAD-Bip” (schizoaffective disorder bipolar type) in the current study. Although the exact symptoms represented by these groups vary somewhat, the general picture is one of significant impairment and substantial clinical severity. Most of the individuals assigned to this class profile in the present study had established diagnoses of either schizoaffective disorder or bipolar I disorder, which aligns with prior studies (Kendler 1998; Boks 2007). The consistent observation of this class profile in multi-diagnosis cohorts suggests that there may be a transdiagnostic group of patients who have severe symptom profiles that more closely resemble each other than less severe forms of the profiles seen in their established diagnostic categories.

Third, four studies observed a latent class symptom profile reminiscent of comorbid schizophrenia and major depression but with overall lower levels of impairment than observed in either classic schizophrenia or the schizobipolar classes. Dubbed “schizodepression” in both Kendler and Peralta and Cuesta, “Cluster 2” (Boks 2007), and Class 5 - SAD-Dep (schizoaffective disorder depressed subtype) in this study, the clinical picture for these classes includes prominent depressive symptoms, but overall, class profile is more similar to schizophrenia than major depression. Psychotic symptoms, chronic illness, and negative symptoms dominate the clinical picture are at comparable levels to the psychotic symptoms reported in the schizophrenia class. This pattern held for all four studies. One notable difference in associated demographics with this symptom profile is that female excess was reported by Boks et al (2007) but not observed in this study. The other studies did not report participant sex proportion by class, so it is unclear how generalizable and replicable this finding is.
